# Microliter whole blood neutrophil assay preserving physiological lifespan and functional heterogeneity

**DOI:** 10.1101/2023.08.28.23294744

**Authors:** Chao Li, Nathan W. Hendrikse, Makenna Mai, Mehtab A. Farooqui, Zach Argall-Knapp, Jun Sung Kim, Emily A. Wheat, Terry Juang

## Abstract

For *in vitro* neutrophil functional assays, neutrophils are typically isolated from whole blood, having the target cells exposed to an artificial microenvironment with altered kinetics. Isolated neutrophils exhibit limited lifespans of only a few hours *ex vivo*, significantly shorter than the 3-5 day lifespan of neutrophils *in vivo*. In addition, due to neutrophils’ inherently high sensitivity, neutrophils removed from whole blood exhibit stochastic non-specific activation that contributes to assay variability. Here we present a method - named “μ-Blood” - that enables functional neutrophil assays using a microliter of unprocessed whole blood. μ-Blood allows multiple phenotypic readouts of neutrophil function (including cell/nucleus morphology, motility, recruitment, and pathogen control). In μ-Blood, neutrophils show sustained migration and limited non-specific activation kinetics (<0.1% non-specific activation) over 3-6 days. In contrast, neutrophils isolated using traditional methods show increased and divergent activation kinetics (10-70% non-specific activation) in only 3 h. Finally, μ-Blood allows the capture and quantitative comparison of distinct neutrophil functional heterogeneity between healthy donors and cancer patients in response to microbial stimuli with the preserved physiological lifespan over 6 days.

## Introduction

Immune functional information ranging from phenotypic level to molecular level has a long history of being studied and used as biomarkers in medicine. Whole blood immune profiling (e.g., flow cytometry, clinical cell counts, and other cellular markers) has been used in patient stratification^1^ and immune signature studies^2–4^. Whole blood immune profiling provides critical data for clinical decision making; however, these assays provide only a snapshot of the chemical and physical markers of the immune cells in whole blood. In comparison, immune cell functional assays - interrogation and quantification of immune cell phenotypic response against stimuli (e.g., drug/pathogen challenge), provide valuable and additional biomarkers on top of the snapshot information from whole blood immune profiling, enabling a more complete set of information for *in vitro* diagnosis (IVD)^5^ and drug/therapy screening^6,7^.

While *in vivo* tests (e.g., mice, zebrafish)^8^ are available and the current gold standard for immune cell functional assays, *in vitro* assays allow improved physical, optical, and biochemical access/challenges and more importantly, the use and interrogation of primary immune cells directly from patients and thus a promising pipeline that leads to personalized/precision medicine^9^. In the *in vitro* regime, various microfluidic systems have been developed to perform whole blood immune profiling (including genomics, transcriptomics, proteomics, and physical markers)^10–12^ and immune cell functional assays (e.g., cell-to-cell, cell-to-pathogen, cell-to-environment)^7^. Despite this progress, the inherently high sensitivity of immune cells (especially neutrophils) and the observer artifacts in an assay still limit the extraction of donor-specific information. In addition, these limitations compound the resolution (e.g., inherent single-cell heterogeneity versus variation caused by artificial environmental factors) and parallel comparison of results from different sources (e.g., experiments, operators, and labs)^13,14^.

In microfluidic immune cell functional assays, the target immune cells are typically captured^12,15–17^ or isolated^18^ from whole blood and then cultured/interrogated in an artificial media (i.e., non-whole blood) microenvironment. These operations expose the sensitive cells to an altered microenvironment^19^ compared to the original whole blood microenvironment that includes autologous signaling molecules^20,21^, other constituent cells [e.g., red blood cells (RBCs)^22^, platelets^23,24^, innate-adaptive crosstalk^25^], and physiopathologically defined oxygen homeostasis/kinetics^26^. Recently, whole blood has been used directly as the assay input in microfluidic immune cell functional assays to improve efficiency, (i.e., sample-to-answer turnaround time) and more importantly to reduce the observer artifacts. However, whole blood still needs to be pre-processed (e.g., RBC lysis or dilution with artificial media and animal serum)^5^ and/or analyzed in/with artificial buffer solutions^12^. Due to the altered and/or loss of the *in vivo* environmental factors, *in vitro* immune cell functional assays still suffer the limited consistency and donor specificity of the information extracted from the assays.

Here we develop a method - named μ-Blood - for neutrophil functional assays with a microliter of unprocessed (i.e., raw) whole blood for both the assay input and through the complete assay, aiming to minimize the observer artifacts and further improve the extraction of donor-specific information and assay consistency. μ-Blood is established on a newly introduced sub-branch in open microfluidics known as Exclusively Liquid Repellency (ELR)-empowered Under-oil Open Microfluidic System (UOMS)^27–33^. μ-Blood allows immune cell functional assays to be performed using and through a micro- or sub-microliter (per testing unit) of unprocessed whole blood and therefore, with the donor-specific whole blood microenvironment preserved. In addition, removal of the whole blood processing steps (e.g., blood cell lysis, blood dilution, cell capture/isolation) reduces the experimental variability and improves the efficiency, consistency, and throughput.

We validate μ-Blood with neutrophils - highly sensitive immune cells - by directly comparing whole blood with isolated neutrophils. The lifespan of isolated neutrophils is short and compounded by non-specific activation (10-70%) - i.e., the spontaneous activation of neutrophils to cell death (including apoptosis and NETosis) due to the altered environment from *in vivo* to *ex vivo* - occurs in only a few hours. By contrast, neutrophils are stable in their whole blood with minimal non-specific activation (<0.1%) for more than 3 days. Using μ-Blood, blood samples from healthy donors and cancer patients are interrogated and compared through 24-h and 6-day assays. The results reveal a distinct neutrophil functional heterogeneity between healthy donors and cancer patients - neutrophils from cancer patients are significantly desensitized compared to the cells from healthy donors. Specifically, cancer patients show: i) upregulated neutrophil count in whole blood [average 4 ± 2 million/mL (cancer) versus average 3 ± 2 million/mL (healthy)]; ii) similar migration speed (average 0.2-0.3 μm/s) from fresh blood in response to fMLP (N-Formylmethionine-leucyl-phenylalanine - a standard neutrophil chemoattractant) at early stage (<3 h) but, significantly lower migration speed (average <0.1 μm/s) in response to live bacteria [*Staphylococcus aureus* (*S. aureus*)] at early stage (<3 h) compared to healthy donors (average 0.3 μm/s); iii) (average 60%) less recruitment at early stage (<3 h) then slowly increased recruitment to a similar level but much lower activation rate [average 13% (cancer) versus average 72% (healthy)] at later stage (∼24 h); and iv) much longer “induction time” [>1 day (cancer) versus <1 h (healthy)] to respond to microbial stimuli.

## Results

### Preservation of the donor-specific whole blood microenvironment - the underlying feature of μ-Blood

The key innovation of the μ-Blood method (Fig. 1) is to not only directly use a microliter (or less) of unprocessed whole blood (i.e., without pre-processing) as the assay input but also perform complete neutrophil functional assays in a whole blood microenvironment. This key enabling feature allows minimized non-specific activation of neutrophils from *in vivo* to *ex vivo* and thus improved extraction of donor-specific information and assay consistency. While whole blood has been reported and used as assay input in *in vitro* neutrophil functional assays, the whole blood either has to be still pre-processed, e.g., diluted with artificial cell culture media, or only used as the assay input. Neutrophils are still removed (e.g., isolated or captured) from whole blood and then interrogated in a non-whole blood microenvironment throughout the readout collection.

**Fig. 1.**
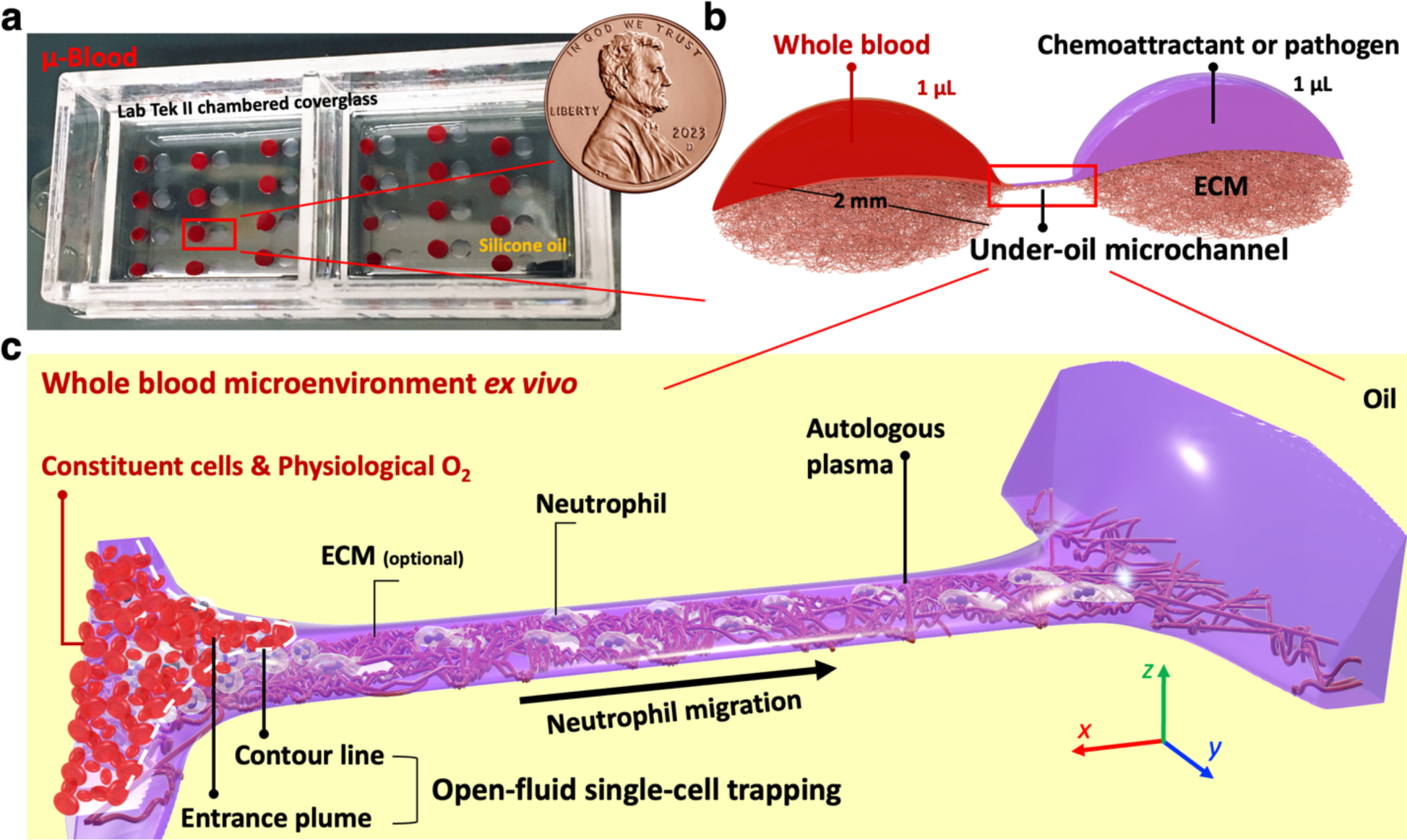
An overview of the μ-Blood system and the whole blood microenvironment. **a**, A camera photo of a μ-Blood device overlaid with silicone oil (5 cSt, 1 mL/well). **b**, A three-dimensional (3D) model that illustrates the geometry of the under-oil microchannel. **c**, A zoomed-in picture of the under-oil microchannel 3D model in (**b**) that shows the fine structures of the whole blood microenvironment and the mechanism of open-fluid single-cell trapping. Blood cells get trapped in the entrance plume defined by the contour line (i.e., a line of equal height) of a given cell size (e.g., RBCs). The space beyond the contour line in the microchannel has a channel height smaller than the defining cell size. Defined by the unique under-oil microenvironment, cells bigger than the channel height can only move into and through a microchannel via active cell migration.

In μ-Blood, the under-oil microchannels with micrometer-scale lateral resolution [with or without extracellular matrix (ECM) coating] (Fig. 1a,b, Section I in SI, Supplementary Fig. 1) can be readily prepared with autologous plasma (separated from the same whole blood sample, Methods) by an operation known as under-oil sweep distribution enabled by Double-ELR^30^, creating a complete whole blood microenvironment *ex vivo* (Fig. 1c). The lateral resolution of surface pattern in ELR-empowered UOMS is defined by the surface patterning method, e.g., photolithography used in this work but not limited to about 30 μm. Defined by the unique geometry of the Double-ELR under-oil microchannels (Fig. 1b,c), a channel height (i.e., the thickness of the media layer or media + ECM layer in a microchannel) less than 2.5 μm - defined by the Laplace pressure equilibrium of the system - can be achieved spontaneously with a much larger channel width at 100 μm (Supplementary Fig. 2a,b). Enabled by the micrometer-scale lateral resolution and the unique geometry of the Double-ELR under-oil microchannels (Fig. 1c), blood cells and bacteria can be reliably and selectively trapped (Supplementary Fig. 2c,d) with free physical access to the samples of interest on a μ-Blood device (Fig. 1a). The blood cells in an under-oil microchannel with single-cell trapping take a monolayer of cells, which is essential for uncompromised optical access to the cells with unprocessed whole blood and *in situ* phenotypic assays (e.g., cell segmentation and tracking). In addition, our real-time oxygen monitoring results showed that whole blood can retain the physiological oxygen concentration (about 5% O_2_ for venous blood) *ex vivo* for nearly 2 days (Supplementary Fig. 3). Together, the preserved whole blood microenvironment including constituent cells, autologous signaling molecules, and physiological O_2_ in μ-Blood provides an exclusive platform to run days-long neutrophil functional assays *ex vivo* with minimized non-specific activation (see detailed results of non-specific activation in the following sections), improved extraction of donor-specific information, and assay consistency.

### Altered neutrophil activation from *in vivo* to *ex vivo*

Immune cells that form the central defense system in our body are extremely sensitive to the environmental factors to which they are exposed. When they get removed from their original niche (e.g., blood, tissue), the immune cells get non-specifically activated without being exposed to/challenged by operator-defined stimuli, simply due to the altered microenvironment. For example, neutrophils sense (“know”) RBCs in blood^22^. If neutrophils get separated and lose the signal from RBCs, they get activated spontaneously. In addition, our studies and some recent works from other researchers showed that immune cells take noticeably altered activation kinetics when kept in whole blood compared to standard culture media usually supplemented with animal serum. Overall, the altered immune cell activation can be attributed to a synergy of physiological factors including autologous signaling molecules, constituent cells, and oxygen homeostasis/kinetics. Running immune cell functional assays in unprocessed whole blood is envisioned as necessary to preserve the physiological kinetics and functions and thus improve the extraction of donor-specific information of the target immune cells. Besides the altered cell activation and loss of donor-specific information during and after transition from *in vivo* to *ex vivo*, the inherent sensitivity of immune cells leads to significant variability/inconsistency from cell isolation (Section II in SI, Supplementary Fig. 4), which not only adds extra operation steps that burden the assay turnaround time/cost/personnel training but also compound the parallel comparison of the results from different sources.

### Comparison of neutrophil non-specific activation between whole blood and isolated neutrophils in different media conditions in μ-Blood

The original driver of developing the μ-Blood method was to minimize the observer artifacts from exposing the target immune cells to a suboptimal or non-physiological assay environment and further improve the extraction of donor-specific information and assay consistency. Our previous study on neutrophil activation *ex vivo* revealed that isolated neutrophils get activated non-specifically with a high variability - for example, in 24 h, 384-well plate, 60K neutrophils in 20 μL RPMI + 10% fetal bovine serum (FBS) + 1% penicillin/streptavidin (Pen/Strep) per well, standard CO_2_ incubator [37 °C, 18.6% O_2_, 5% CO_2_, 95% relative humidity (RH)]; and much more quickly in 12 h, 384-well plate, 60K neutrophils in 20 μL RPMI + 10% FBS + 1% Pen/Strep per well, onstage incubator (37 °C, 21% O_2_, 5% CO_2_, 95% RH). The isolated neutrophils get removed from their original whole blood environment and then cultured/interrogated in an artificial cell culture media with altered nutrient level and signaling molecules (e.g., autologous plasma versus animal serum), constituent cells, and oxygen availability [e.g., 5% O_2_ in venous blood, 13% O_2_ in arterial blood, versus 21% O_2_ in air, 18.6% O_2_ in standard cell incubator]. By contrast, neutrophils remain stable (i.e., non-activated) in whole blood *ex vivo* for at least 2 to 3 days. In whole blood, even if already drawn out of the body, neutrophils are still exposed to autologous signaling molecules and constituent cells. Moreover, the results from the real-time oxygen monitoring showed that whole blood can retain 5% O_2_ in venous blood outside of the body in air for up to 2 days (Supplementary Fig. 3). These pilot studies together explain the distinct neutrophil activation kinetics in whole blood compared to an artificial cell culture/assay environment.

Here, we directly compare neutrophil non-specific activation between unprocessed whole blood and isolated neutrophils on a μ-Blood device against different culture media conditions - 100% autologous platelet-free plasma (A-PFP), 100% heterologous (i.e., donor-mismatched, prostate cancer in this experiment) PFP (H-PFP), 100% FBS, RPMI (the standard basal media for culturing suspension cells) + 20% A-PFP, RPMI + 20% H-PFP, and RPMI + 20% FBS (Fig. 2a-c). To keep a donor-specific autologous microenvironment, we used microchannels prepared with PFP, without ECM coating, and with double-oil overlay for long-term (days) assays (Section III in SI, Supplementary Fig. 5).

**Fig. 2.**
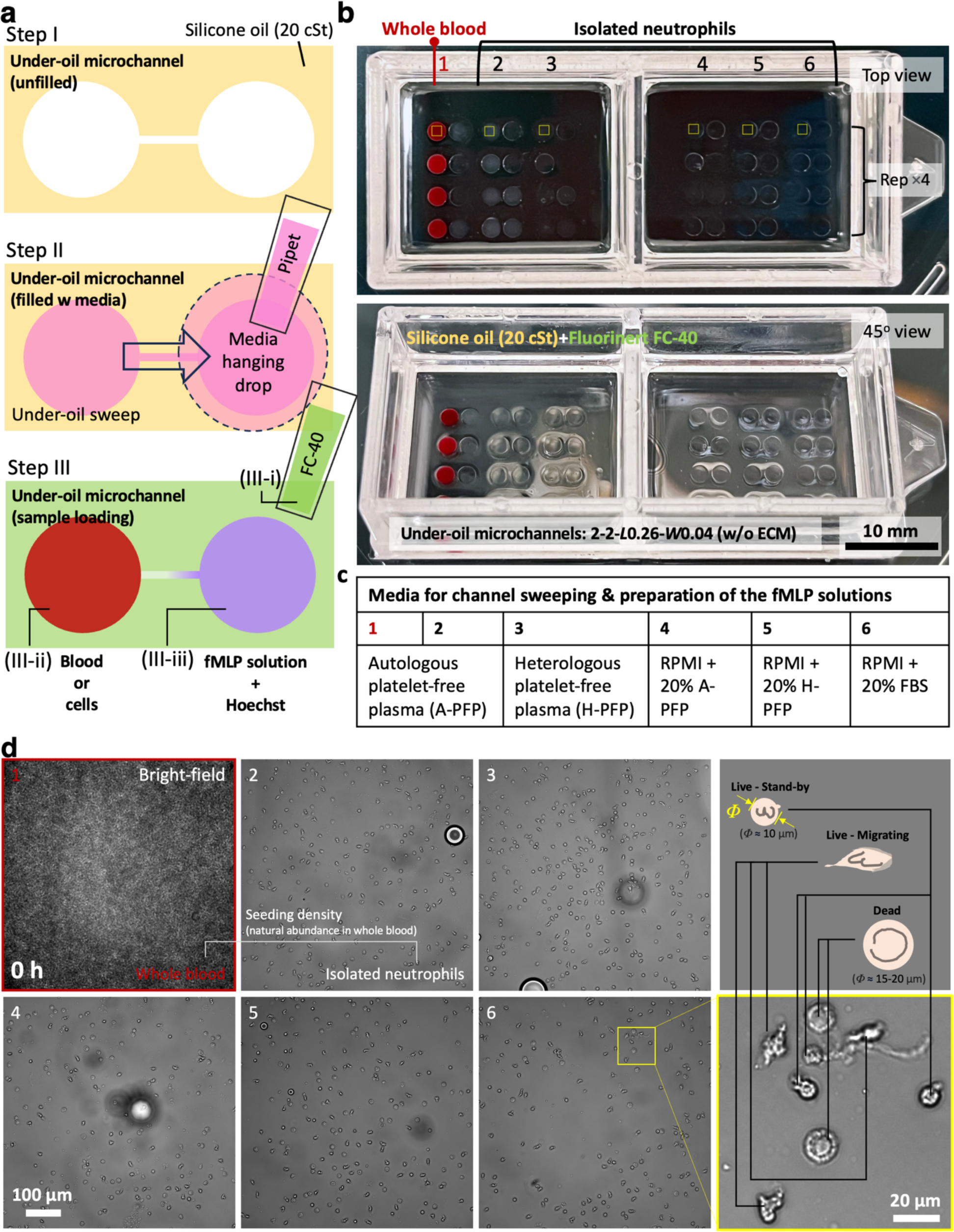
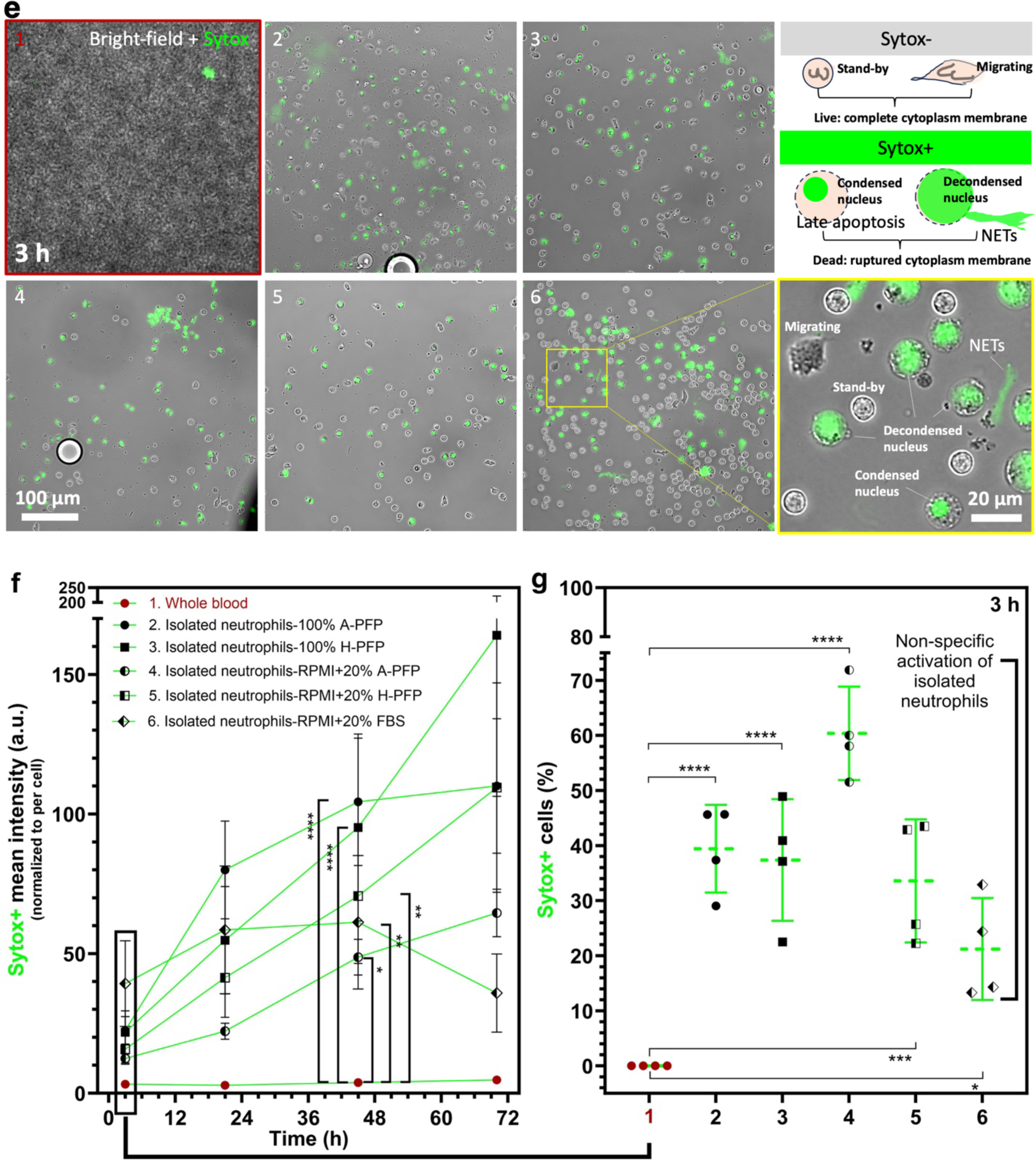
Side-by-side comparison of neutrophil non-specific activation (i.e., baseline cell death without operator-defined stimuli) between whole blood and isolated neutrophils for 3 days in μ-Blood. **a**, Schematic showing the preparation steps of an under-oil microchannel (without ECM coating) and sample loading with double-oil [i.e., silicone oil (20 cSt) or “SO20” + fluorinated oil (Fluorinert FC-40) or “FC-40”] overlay (Section V in SI, Supplementary Fig. 7). The spots and channel are not in scale for illustration. **b**, Camera photos showing the layout of the μ-Blood device. The yellow boxes indicate the region of interest (ROIs) in (**d**) and (**e**). **c**, The culture media conditions (1 to 6) for channel sweeping and preparation of the fMLP solutions. **d**, Bright-field images showing the optical access and cell seeding density of isolated neutrophils at 0 h (i.e., right afte cell seeding) on the μ-Blood device. The neutrophil seeding density is defined by the natural abundance in the whole blood sample (i.e., 2× of neutrophil count from EasySep isolation with an average yield of 50%). (Callouts) Neutrophil stages identified by bright-field cell morphology. **e**, Non-specific activation (Sytox+: late apoptosis and NETosis) of neutrophils at 3 h on μ-Blood device. (Callouts) Neutrophil stages identified by bright-field and Sytox (a cytoplasmic membrane-impermeable DNA stain) cell morphology. **f**, Comparison of non-specific activation of neutrophils between whole blood and isolated neutrophils up to 3 days on the μ-Blood device. The y axis is the mean fluorescence intensity from the Sytox channel normalized to the total number of cells in the field of view in each condition. **g**, Sytox+ cell percentage (i.e., Sytox+ cell count/total cell count × 100%) at 3 h after cell seeding. The non-specific activation of isolated neutrophil on the μ-Blood device ranges from about 10% to 70% in different culture media conditions. Error bars are mean ± S.D. from ≥3 replicates. \**P* ≤ 0.05, \*\**P* ≤ 0.01, \*\*\**P* ≤ 0.001, and \*\*\*\**P* ≤ 0.0001. “ns” represents “not significant”.

On the μ-Blood device, the results showed that isolated neutrophils got non-specifically activated ranging from 10% to 70% in only 3 h (under-oil 2 mm spot, 1500 cells/μL per spot, onstage incubator). By contrast, neutrophils in whole blood on the same device remained stable with minimal (<0.1%) non-specific activation for more than 3 days (under-oil 2 mm spot, 1 μL whole blood per spot, onstage incubator) (Fig. 2d-g). These results are consistent with our previous study of non-specific activation of isolated neutrophils *ex vivo* (Supplementary Fig.4d). The altered and stochastic neutrophil activation in an artificial culture/assay environment pinpoints the necessity of using unprocessed whole blood and autologous plasma to perform immune cell functional assays. μ-Blood can be performed with or without ECM (Fig. 1c, Supplementary Fig. 2c,d). If ECM is used, it is worth noting that the source of ECM is limited to animals (e.g., murine, bovine). In ECM-free μ-Blood, all the reagents involved are from the same whole blood sample for a complete, donor-specific whole blood microenvironment (Fig. 1c).

### Translational application - donor-specific neutrophil functional heterogeneity

In this section, we demonstrate the translational application of ECM-free μ-Blood with clinical blood samples. We first compared neutrophil non-specific activation between 10 healthy (self-reported) donors and 26 cancer patients (Section IV in SI, Supplementary Fig. 6, Supplementary Fig. 7, Supplementary Fig. 8). The results showed that neutrophils from healthy donors were more sensitive/responsive to the altered environmental factors from *in vivo* to *ex vivo* compared to cancer patients. Neutrophils from healthy donors featured a more N1-like (i.e., pro-inflammatory) phenotype and in contrast, the desensitized neutrophils from cancer patients displayed a more N2-like (i.e., anti-inflammatory) phenotype (Supplementary Fig. 7). Here, we further characterize the donor-specific neutrophil functional heterogeneity by comparing the recruitment and activation of neutrophils in response to fMLP (Fig. 3) and live bacteria (*S. aureus*) (Fig. 4) in μ-Blood. In these experiments, we used venipuncture blood [with anticoagulant citrate dextrose (ACD) solution, see Section V in SI for the influence of anticoagulants, Supplementary Fig. 9, Supplementary Movie 1] from healthy donors and cancer patients. For the fMLP experiments, we chose the microchannels 2-2-*L*0.26-*W*0.04 (w/o ECM) (see Section VI in SI for the influence of channel dimensions) prepared with 100% A-PFP that allow i) a relatively narrow migration front in the microchannel with 1-3 cells in parallel (Supplementary Movie 2-2), and ii) a relatively small neutrophil count (<50) in/through the channel to facilitate cell count/tracking (Supplementary Fig. 10). For the bacteria experiments, we chose the microchannels 2-2-*L*0.5-*W*0.1 (w/o ECM) prepared with 100% A-PFP that allow i) a broader migration front in the microchannel with 3-8 cells in parallel (Supplementary Movie 2-1, Supplementary Movie 2-3), and ii) neutrophil-pathogen interaction in an A-PFP environment with RBCs (Supplementary Fig. 2c, Supplementary Movie 2-3).

**Fig. 3.**
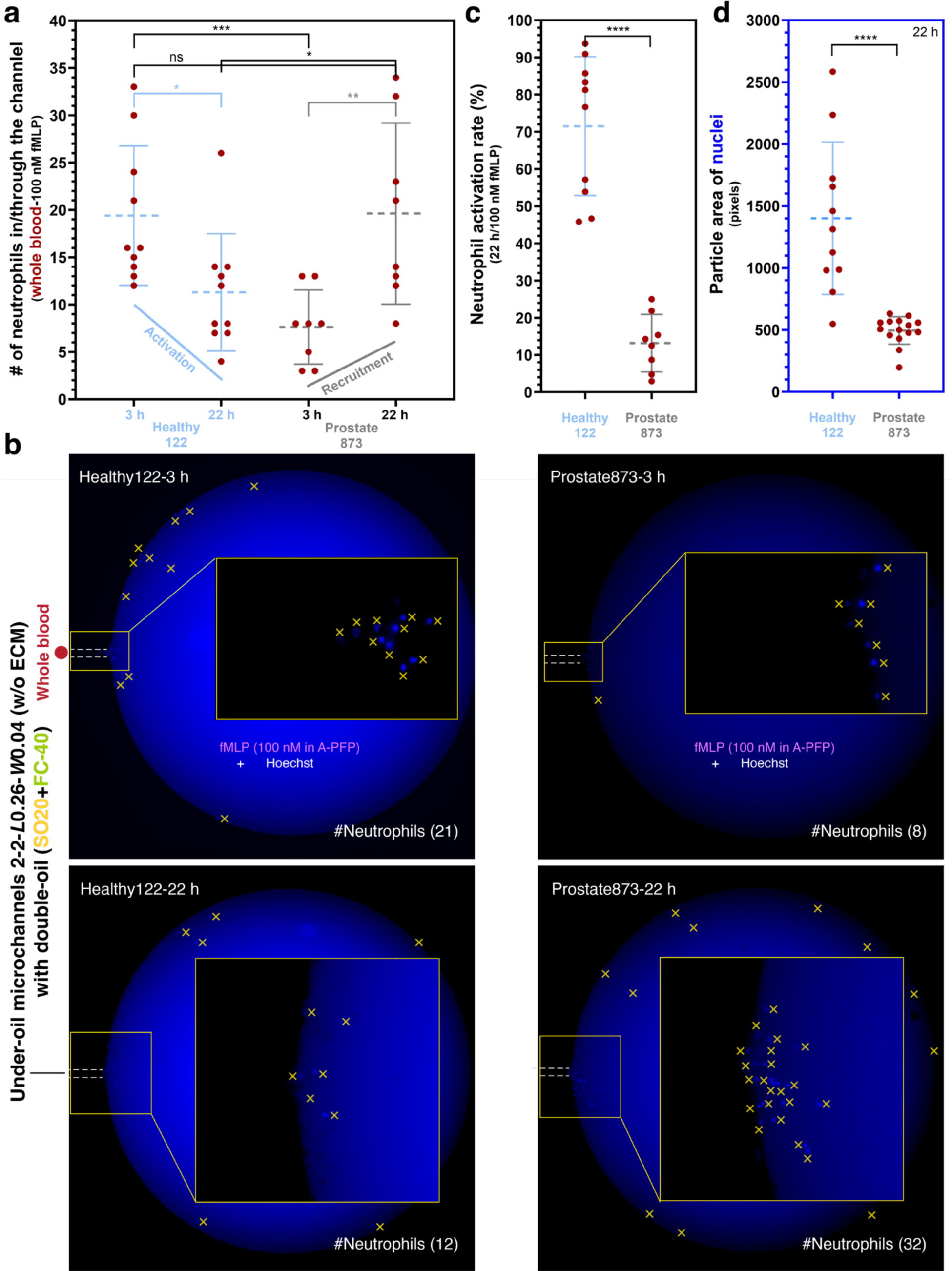
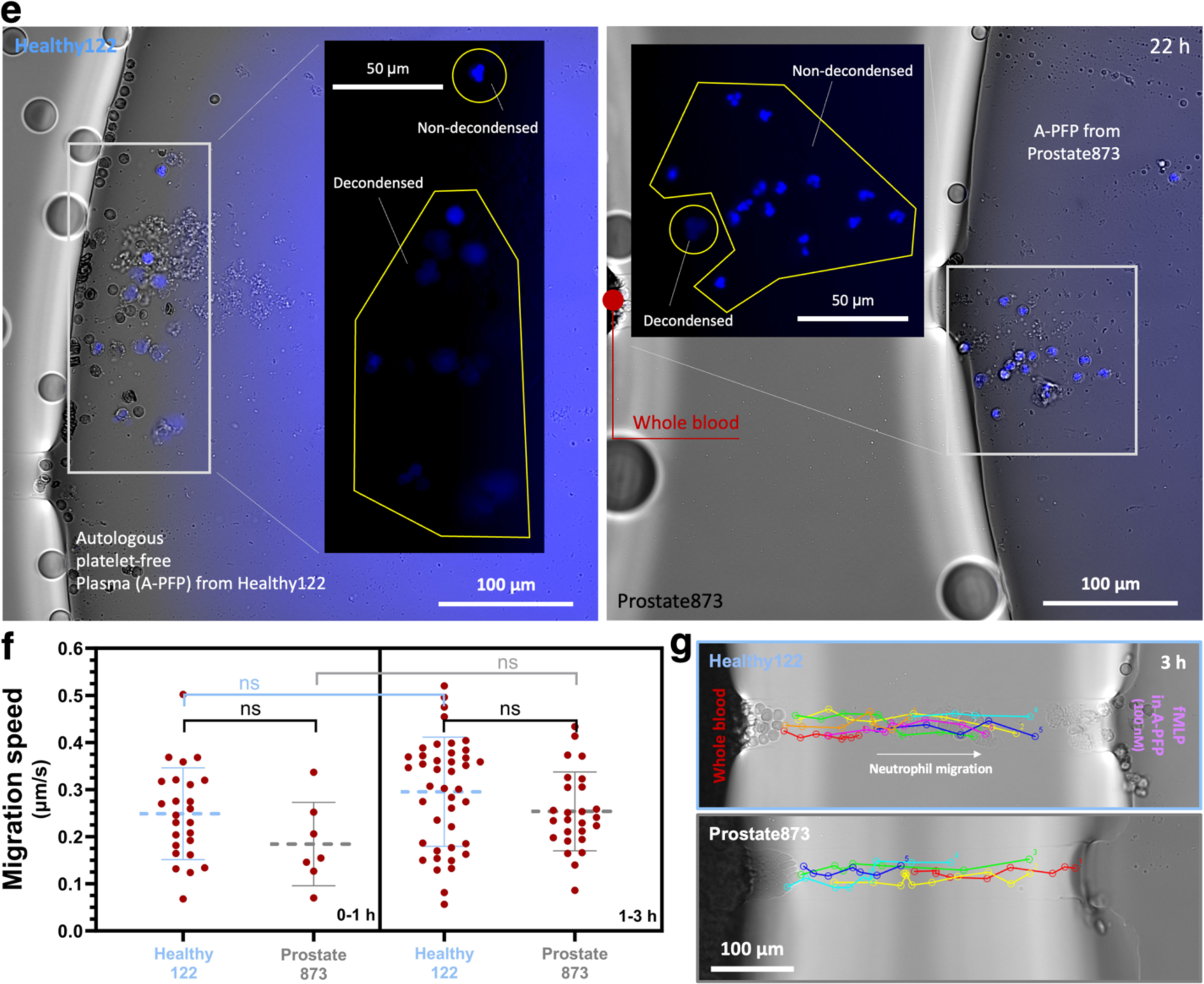
Comparison of neutrophil function including recruitment and activation against fMLP between a healthy donor and a cancer patient for 24 h in μ-Blood. **a**, Neutrophil recruitment from 1 μL of unprocessed whole blood at 3 h and 22 h against 100 nM fMLP. The microchannels [2-2-*L*0.26-*W*0.04 (w/o ECM)] were prepared with A-PFP and double-oil overlay by under-oil sweep (Fig. 4a). Prostate873 neutrophils showed (1-7/19) × 100% = 63% less recruitment at 3 h and then slowly increased to a similar level at 22 h compared to Healthy122 neutrophils. The decrease of neutrophil count of Healthy122 at 22 h compared to 3 h is attributed to the high activation rate of neutrophils shown in (**c**). **b**, Representative microscopic images (Hoechst fluorescence from nuclei) of (**a**). The yellow cross signs indicate the neutrophils found on the fMLP spots from migration. It is worth noting that Hoechst (1:200 dilution) shows a high background when used in human plasma but nearly no background in FBS. (Insets) Zoomed-in images of neutrophil nuclei. **c**, Neutrophil activation rate (%) - the number of neutrophils with decondensed nuclei (i.e., activated)/the total number of neutrophils in/through the channel × 100%. Prostate873 neutrophils showed significantly lower activation rate (13%) compared to Healthy122 neutrophils (72%) at 22 h. It needs to be noted that the total number at 3 h was used to calculate the activation rate of Healthy122 neutrophils at 22 h to compensate for the drop of neutrophil count. **d**, Particle area of nuclei at 22 h. The mean particle area of nuclei from Healthy122 neutrophils is 1400 pixels/500 pixels = 2.8 times of that from Prostate873 neutrophils due to decondensation of nuclei. **e**, Representative microscopic images (composite: bright-field + Hoechst fluorescence of nuclei) show the decondensed and non-decondensed nuclei (Fig. 4e) at 22 h on the fMLP spots. **f**, Migration speed of Healthy122 and Prostate873 neutrophils in 0-1 h and 1-3 h. **g**, Representative cell migration tracks of (**f**). Error bars are mean ± S.D. from ≥3 replicates. \**P* ≤ 0.05, \*\**P* ≤ 0.01, \*\*\**P* ≤ 0.001, and \*\*\*\**P* ≤ 0.0001. “ns” represents “not significant”.

**Fig. 4.**
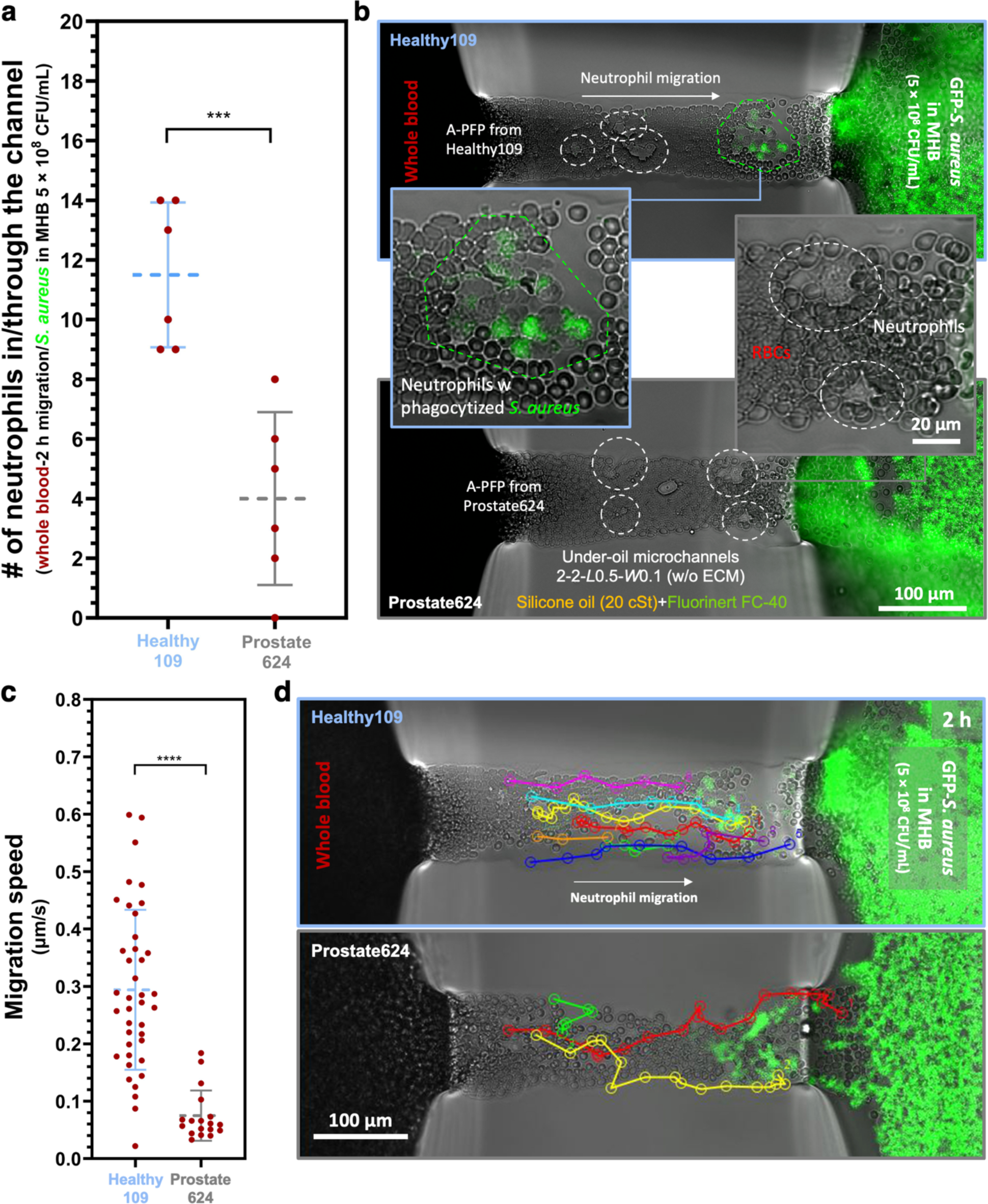
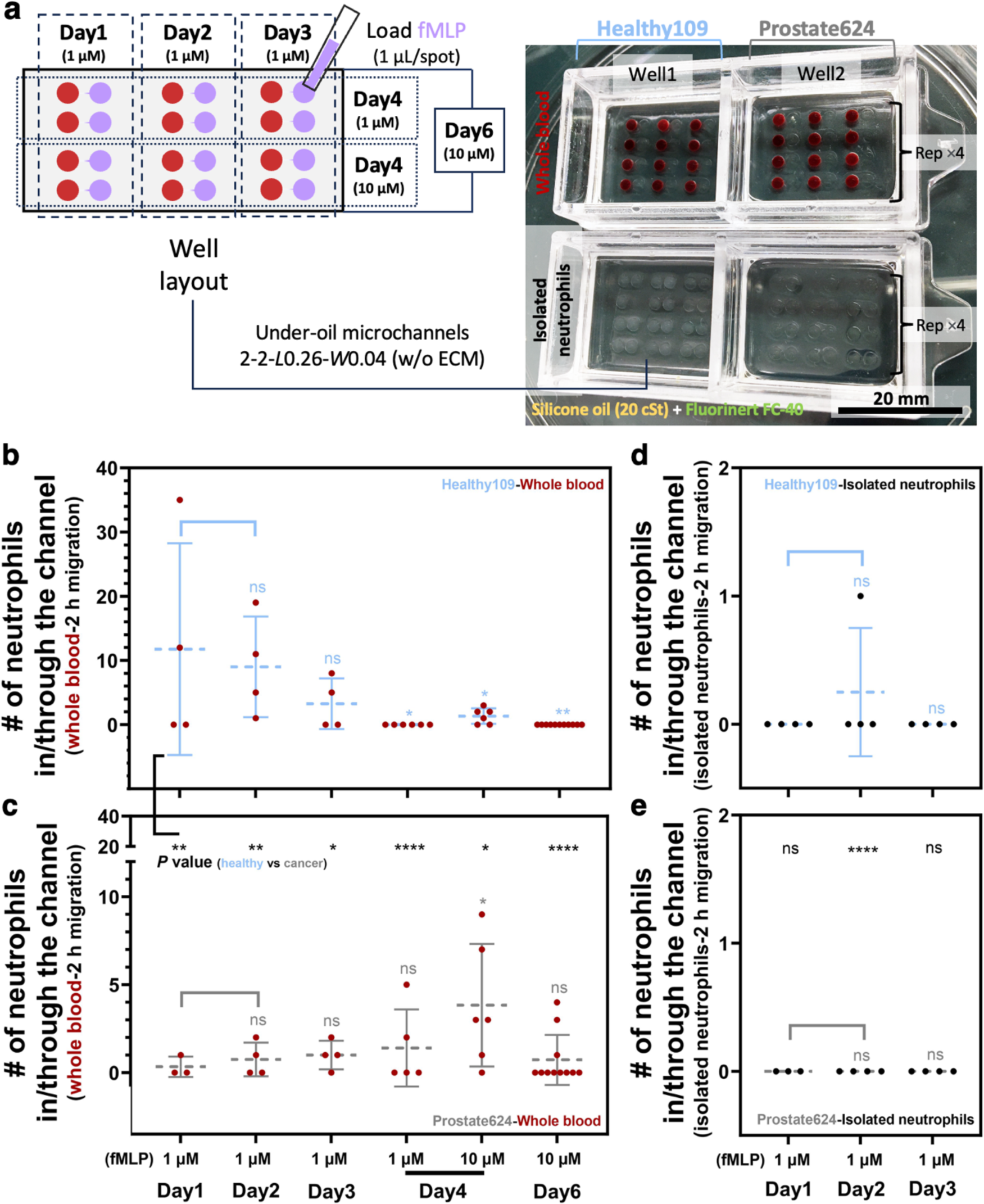
Comparison of neutrophil response to live bacteria (*S. aureus*) between a healthy donor and a cancer patient for 2 h in μ-Blood. **a**, Neutrophil recruitment from 1 μL of unprocessed whole blood in 2 h against *S. aureus* [GFP-labeled, 5 × 10^8^ CFU/mL in Mueller Hinton Broth (MHB)]. The microchannels 2-2-*L*0.5-*W*0.1 (w/o ECM) were prepared with A-PFP and double-oil overlay by under-oil sweep (Fig. 4a). Prostate624 neutrophils showed (1-4/11.5) × 100% = 65% less recruitment at 2 h compared to Healthy109 neutrophils. **b**, Representative microscopic images (composite: bright-field + GFP fluorescence from *S. aureus*) of (**a**). The responsive neutrophils migrating into the channels are marked out by the dashed line boxes. The green dashed-line box highlights the phagocytosis of GFP-*S. aureus* by neutrophils from the healthy donor. **c**, Migration speed and **d**, representative cell migration tracks of (**c**). Healthy109 neutrophils showed significantly higher migration speed (average 0.28 μm/s) compared to Prostate624 neutrophils (average 0.05 μm/s). Error bars are mean ± S.D. from ≥3 replicates. \**P* ≤ 0.05, \*\**P* ≤ 0.01, \*\*\**P* ≤ 0.001, and \*\*\*\**P* ≤ 0.0001. “ns” represents “not significant”.

Whole blood samples from a healthy donor (Healthy122) and a cancer patient (Prostate873) were compared at an early stage of 2-3 h and a later stage at around 24 h. In response to 100 nM fMLP in A-PFP at 3 h, Healthy122 showed a much higher recruitment of neutrophils (average 19 neutrophils/channel) compared to Prostate873 (average 7 neutrophils/channel), accounting for average 60% less requirement of Prostate873 neutrophils at the early stage (Fig. 3a,b). After the first 3-h timelapse on a microscope in an onstage incubator, the μ-Blood device was kept in a standard CO_2_ incubator for the later time point. At 22 h, Prostate873 neutrophils showed a significantly increased recruitment to a level comparable to Healthy122 neutrophils at 3 h. In comparison, the number of neutrophils on the fMLP spot from Healthy122 decreased significantly from average 19 neutrophils/channel at 3 h to average 11 neutrophils/channel at 22 h (Fig. 3a,b). We further quantified the neutrophil activation rate at 22 h by segmenting cells on the fMLP spots with the morphology of nuclei - decondensed^34^ (i.e., activated) (Fig. 2e, Fig. 3c-e). The results showed that the activation rate of Healthy122 neutrophils (average 72%) was much higher than Prostate873 neutrophils (average 13%). The quick and strong activation and decondensation of nuclei account for the drop of neutrophil count on Healthy122 in the 24-h assay (Fig. 3a,b).

In response to a given stimulus, a low recruitment of neutrophils can be attributed to #1) down-regulated neutrophil count in whole blood (i.e., fewer neutrophils per μL of whole blood), #2) reduced motility, and #3) other donor-specific heterogeneities that lead to reduced responsiveness or longer induction time to respond to stimuli. Our results of neutrophil count from direct whole blood neutrophil isolation showed that cancer patients typically come with a higher neutrophil count (average 4000 cells/μL, 26 patients) compared to healthy donors (average 3000 cells/μL, 10 donors) (Supplementary Fig. 6c). Further, the migration speed analysis (Fig. 3f,g, Supplementary Movie 3) showed that Healthy122 and Prostate873 neutrophils came with similar motility (average 0.2-0.3 μm/s) in response to fMLP in 3 h. Prostate873 neutrophils seemed to be slower in migration speed but had no statistical significance compared to Healthy122 neutrophils. Together, these fMLP-test results (Fig. 3) indicate that the less recruitment of neutrophils from cancer blood is related to a reduced responsiveness to stimuli, which is likely to be caused by other donor-specific heterogeneities rather than neutrophil count in whole blood or motility.

fMLP is a bacteria peptide and a standard chemoattractant for studying/quantifying neutrophil chemotaxis. Live bacteria secrete many chemicals that may lead to a different neutrophil response compared to pure fMLP. Next, we switched to live bacteria (GFP-labeled *S. aureus*) to test neutrophil response and pathogen control in μ-Blood with whole blood and an A-PFP microchannel with autologous RBCs (Fig. 4a,b). Here, we also compare a healthy donor (Healthy109) and a cancer patient (Prostate624). The results showed that Healthy109 neutrophils came with a significantly higher recruitment toward *S. aureus* at 2 h compared to Prostate624 neutrophils (Fig. 4a), consistent with the recruitment results in response to fMLP (Fig. 3a). In the 2-2-*L*0.5-*W*0.1 (w/o ECM) microchannels with a channel height of about 2.3 μm (Supplementary Fig. 2b, inset), neutrophils were able to smoothly navigate through the RBC monolayer in the microchannel (Supplementary Fig. 2c) and phagocytize bacteria quickly (in <2 min) after the encounter (Fig. 4b). The phagocytosis process was readily identified by internalization of GFP-*S. aureus* blobs into neutrophils and then getting carried over with neutrophil movement (Supplementary Movie 4). The phagocytosis events were primarily observed with Healthy109 neutrophils in the microchannels. The apparently reduced phagocytosis from Prostate624 neutrophils could be a direct result of less recruitment and thus lower density of neutrophils in the microchannels, while the possibility of reduced phagocytosis ability is not ruled out and will be further investigated. Essentially different from fMLP, live bacteria proliferate and modify the immune-pathogen microenvironment over time dynamically. Longer (>2 h) neutrophil-*S. aureus* interactions were studied in μ-Blood but not included in this report.

Further, the migration speed analysis (Fig. 4c,d) revealed that Prostate624 neutrophils were able to respond and migrate toward *S. aureus* through a monolayer of RBCs but in a much slower migration speed (average 0.05 μm/s) up to about 0.2 μm/s. By contrast, Healthy109 neutrophils, in the parallel testing environment (i.e., in response to *S. aureus* and through RBCs), responded much quicker and stronger with a high migration speed (average 0.28 μm/s) up to about 0.6 μm/s. This distinct difference of migration speed contributes to the less recruitment toward *S. aureus* from Prostate624 compared to Healthy109 (Fig. 4a). These bacteria-test results further indicate the reduced responsiveness of neutrophils from cancer patients in response to microbial stimuli.

### Long-term (days) neutrophil kinetics between healthy-donor and cancer-patient neutrophils

The role of innate immune (e.g., neutrophils, macrophages) in cancer diseases and treatments continues to gain attention^35,36^. So far most of the neutrophil functional assays were performed on animal models^37,38^ as neutrophil is notorious with its inherently high sensitivity and difficulty to manipulate *ex vivo*^39^. The reported *in vitro* neutrophil functional assays were typically performed with a pure isolate of neutrophils or pre-processed whole blood (RBC lysis or dilution with artificial media and animal serum) with a standard assay time window for only a few hours. Long-term (days) *in vitro* neutrophil functional assays were rarely explored/performed due to the challenges from both biology (i.e., neutrophil sensitivity and non-specific activation) and platform engineering. The high neutrophil sensitivity leads to stochastic, non-specific activation after the cells are removed from whole blood (Fig. 2, Section II in SI, Supplementary Fig. 4). On top of this biological challenge and as aforementioned in the first section of Results, to perform *in vitro* neutrophil functional assays in microfluidics, microchannels that support single-cell trapping are required. The media volume in single-cell trapping microchannels falls into the nanoliter to picoliter range (Section VI in SI, Supplementary Table 1), which is vulnerable to environmental/operational factors such as media loss via evaporation (Section III in SI, Supplementary Fig. 5a) and bubble/particle clogging with the classic closed-channel design.

In the previous sections, we already demonstrated the applicability of μ-Blood to perform standard (i.e., hours) *in vitro* neutrophil functional assays using a microliter of unprocessed whole blood. To run long-term (days) *in vitro* neutrophil functional assays, we introduced a double-oil overlay method (Section III in SI, Supplementary Fig. 5b) to retain hydration of the single-cell trapping microchannels under oil for days (tested up but not limited to 6 days).

Here, we prepared two μ-Blood devices to compare whole blood with isolated neutrophils and the possible long-term neutrophil functional heterogeneity between a healthy donor (Healthy109) and a cancer patient (Prostate624) (Fig. 5a). With the experimental design, we added fresh fMLP solution (1 μM in A-PFP) to a subset of the microchannels (×4 replicates per condition) through 3 days and then added two different fMLP concentrations (1 μM and 10 μM in A-PFP) on Day 4 to half of the Day-1-to-Day-3 microchannels. At last, on Day 6, fMLP at 10 μM in A-PFP was added to each well for the end point. Through a 6-day assay in whole blood and the A-PFP environment, we successfully extracted the days-long neutrophil kinetics information specific to the donor conditions in μ-Blood (Fig. 5b,c,f-h). By contrast, the control group with isolated neutrophils provided little information that reflects donor-specific heterogeneity (Fig. 5d,e).

**Fig. 5.**
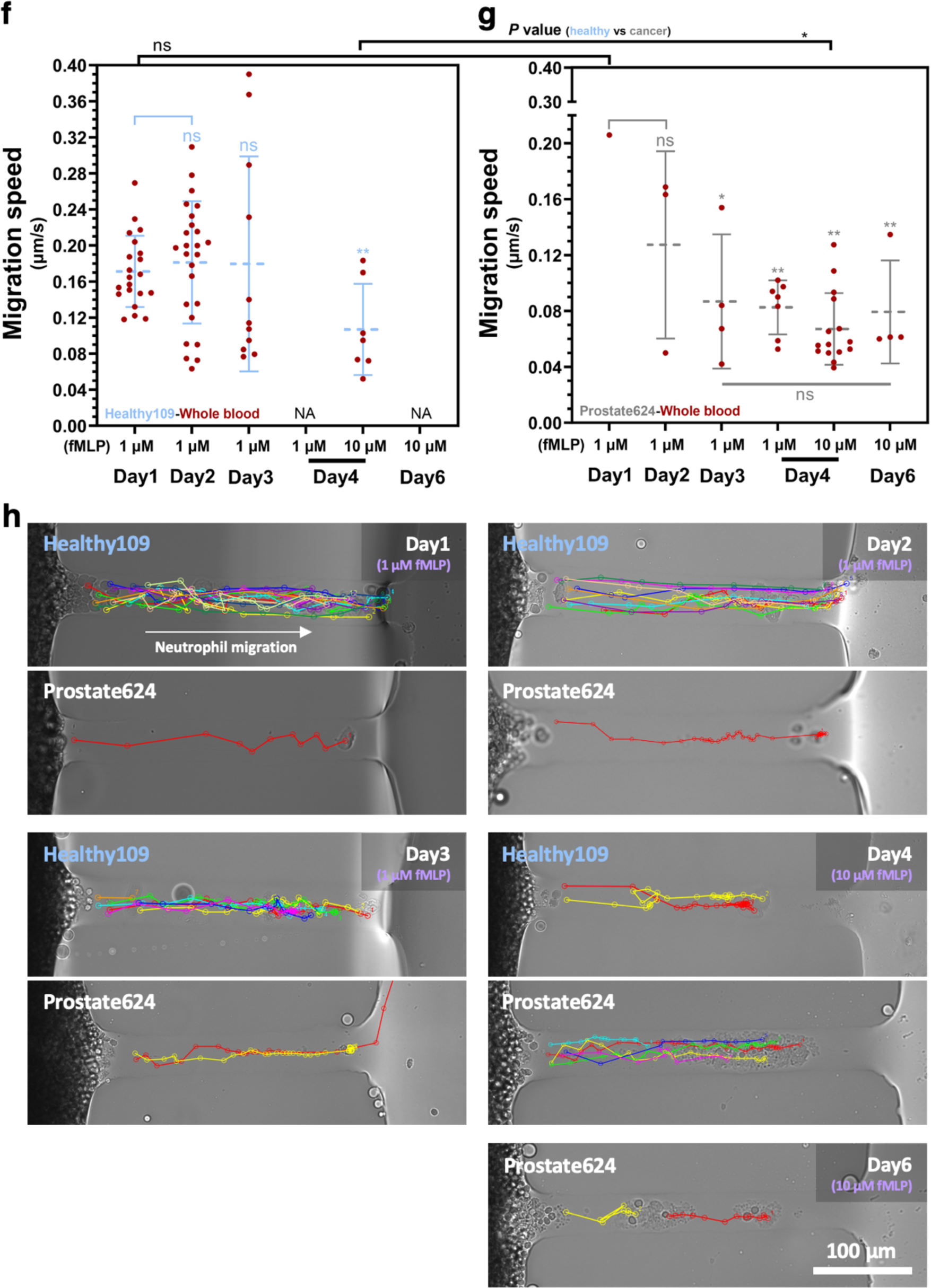
Comparison of neutrophil recruitment against fMLP between a healthy donor and a cancer patient for 6 days in μ-Blood. **a**, A schematic and a camera photo showing the layout of two μ-Blood devices (Device 1 - whole blood; Device 2 - isolated neutrophils; Well 1 - Healthy109; Well 2 - Prostate624). Fresh fMLP solution in A-PFP was loaded to the device through 6 days. The devices were imaged in a 2-h timelapse each day in an onstage incubator. After imaging, the devices were kept in a standard CO2 incubator. **b**, (Healthy109) and **c**, (Prostate624) Neutrophil recruitment from 1 μL of unprocessed whole blood in 2 h. **d**, (Healthy109) and **e**, (Prostate624) Neutrophil recruitment from isolated neutrophils in 2 h. Migration speed of **f**, Healthy109 neutrophils and **g**, Prostate624 neutrophils in 6 days. **h**, Representative cell migration tracks (bright-field) of (**f,g**). Error bars are mean ± S.D. from ≥3 replicates. \**P* ≤ 0.05, \*\**P* ≤ 0.01, \*\*\**P* ≤ 0.001, and \*\*\*\**P* ≤ 0.0001. “ns” represents “not significant”.

Similar to the neutrophil recruitment results in the 24-h fMLP test (Healthy122 versus Prostate873) (Fig. 3) and the 2-h bacteria test (Healthy109 versus Prostate624) (Fig. 4), Healthy109 neutrophils showed a quick and strong response (average 12 neutrophils/channel up to 35) in 2 h on Day 1 right after adding fMLP (Fig. 5b, Supplementary Movie 5); however, almost no recruitment (0-1 neutrophil/channel) was observed from Prostate624 neutrophils (Fig. 5c). This recruitment differential was maintained through 3 days followed by an opposite trend of kinetics in neutrophil recruitment - Healthy109 decreasing and Prostate624 slightly increasing over time (Fig. 5b,c). On Day 4, the recruitment differential was flipped - Healthy109 (average 0 neutrophil/channel in response to 1 μM fMLP, 0-3 neutrophils/channel in response to 10 μM) versus Prostate624 (0-5 neutrophils/channel in response to 1 μM fMLP, 0-9 neutrophils/channel in response to 10 μM). Further, on Day 6, Healthy109 showed no neutrophil recruitment in response to 10 μM. In comparison, Prostate624 still showed a recruitment at the level of 0-4 neutrophils/channel on Day 6. The results from parallelly isolated neutrophils failed to capture the long-term neutrophil kinetics and donor-specific functional heterogeneity (Fig. 5d,e) due to the quick/stochastic non-specific activation (Fig. 2f,g).

A further migration speed analysis revealed that cancer-patient neutrophils came with similar migration speed (average 0.2 μm/s) compared to healthy-donor neutrophils from fresh blood at early stage (Fig. 5f-h, Supplementary Movie 5). In addition, healthy-donor neutrophils retained nearly constant high migration speed (average 0.2 μm/s) through 3 days, followed by a significantly decreased migration speed down to average 0.1 μm/s (Fig. 5f) and an intuitively decreased recruitment (Fig. 5b). By contrast, cancer-patient neutrophils showed reduced migration speed (down to average 0.1 μm/s) starting at relatively early stage (Day 2 to Day 3) (Fig. 5g) but counter-intuitively increased recruitment over time (Fig. 5c). This opposite trend between migration speed and recruitment observed on cancer-patient neutrophils, again, indicates a reduced responsiveness apparently characterized as a significantly increased induction time in response to microbial stimuli. Our hypothesis on the reduced responsiveness of cancer-patient neutrophils is that cancer-patient neutrophils could be systematically desensitized and primed toward more anti-inflammatory (N2-like), showing reduced responsiveness and extended induction time against stimuli that normally trigger quick and strong pro-inflammatory (N1-like) responses on healthy-donor neutrophils (Section IV in SI, Supplementary Fig. 6, Supplementary Fig. 7).

## Discussion

Immune cells, especially neutrophils, are amongst the most sensitive cell types due to their natural function as the first line of response/defense against an altered/abnormal microenvironment (e.g., damaged tissues, infections). During the transition from *in vivo* to *ex vivo* and the following interrogation, the altered environmental factors (including temperature, nutrients/vital gas level and kinetics, signaling molecules, constituent cells, and mechanical cues) are enough to activate neutrophils non-specifically and randomly without operator-defined stimuli. In such cases, it is very challenging to extract the donor-specific information from the compromised immune cells. Moreover, the non-specific activation of immune cells *ex vivo* inflicts limited assay consistency.

The design philosophy of the μ-Blood method is to preserve and extract a more complete set of donor-specific information over a physiologically relevant period (e.g., the *in vivo* lifespan of neutrophils for 3-5 days) by keeping and interrogating the target immune cells in a complete whole blood microenvironment. Various microfluidic systems (predominantly closed-chamber/channel systems) have been used to perform *in vitro* immune cell functional assays with improved control and thus relevance of the interrogation microenvironments compared to *in vivo*. Specifically, the use of whole blood as the assay input (versus isolated cells) represents a milestone in this direction as it streamlines the workflow by removing most of the blood cell isolation steps and more importantly, reduces the operator-derived inconsistencies. The latest whole blood-based immune cell functional assay uses whole blood but still diluted in a cell culture media supplemented with animal serum, which makes it suboptimal compared to unprocessed whole blood. In addition, the closed-channel configuration makes it difficult to have free, nondestructive physical access to the single cells of interest on the device for off-chip downstream analyses, e.g., single-cell sequencing and heterogeneity assay.

Different from the closed system-based immune cell functional assays, the proposed μ-Blood method in this report was developed based on ELR-empowered UOMS - a newly introduced sub-branch in open microfluidics with the state-of-the-art lateral resolution (i.e., micrometer scale versus millimeter scale) and open-fluid controls (including open-fluid single-cell trapping, improved flow range, and on-demand open-fluid valves). Directed by the design philosophy aforementioned, μ-Blood uses a small volume (≤1 μL per testing unit) of unprocessed/raw whole blood as the assay input and autologous plasma to prime the microchannels, ensuring a complete whole blood microenvironment through the assays *ex vivo*. Multiple phenotypic readouts can be continuously extracted through a time course or timelapse for both short-time (in 24 h) and long-time (days-long) assays. Moreover, the open-system configuration of μ-Blood allows the operators to collect single cells of interest anytime and anywhere seamlessly and selectively with minimal system disturbance during and after an assay, e.g., using a single-cell aspirator directly on the device under the microscope, which allows single-cell phenotypic screening - a highly desired function in *in vitro* immune cell functional assays for studying inherent immune heterogeneity at single-cell resolution.

The follow-up directions that aim to incubate μ-Blood to a mature, commercially available immune cell functional assay platform include: i) development of numerical models^32^ to predict the fluid dynamics and mass transport in the under-oil microchannels, especially with complicated/customized patterns; ii) development of an onstage, automated single-cell manipulator^29^ for high-throughput sample loading and collection; iii) integration with multiple optical spectroscopy (including multiphoton, Raman, IR) for label-free, *in situ*, single-cell biochemical analyses^40^; and iv) development of machine learning algorithms for real-time single-cell phenotype segmentation (including cell/nucleus morphology, pathogen control, and degradation of ECM) and cell tracking^33^.

In translational research future studies using μ-Blood, broader diseases including multisystem inflammatory syndrome (MIS), autoimmune diseases, and infectious diseases will be investigated with a large (>50) donor pool. Specifically, innate-adaptive immune crosstalk (e.g., neutrophil-T cell, neutrophil-monocyte/macrophage), cancer-immune-microorganism interkingdom interactions, and antimicrobial susceptibility testing can be explored in the donor-specific whole blood microenvironment. We envision μ-Blood (especially in its fully automated version) can be directly used in hospitals/clinics for immune cell functional screening with improved preservation and extraction of donor-specific information and assay efficiency, consistency, and throughput. Especially, μ-Blood provides an optimal solution where access to a large volume of whole blood sample is limited or impossible, e.g., the newborns. μ-Blood also provides an easy-to-adopt, easy-to-use, and multi-functional platform that better supports the studies in fundamental immunology, especially where environmental factors play a critical role on cell functions at single-cell level.

## Methods

### Preparation of ELR-empowered UOMS devices

Step #1) Fabrication of PDMS silane-grafted surface. Chambered coverglass [Nunc Lab-Tek-II, 2 well (155379), #1.5 borosilicate glass bottom, 0.13 to 0.17 mm thick; Thermo Fisher Scientific] (Fig. 1a) was treated first with O_2_ plasma (Diener Electronic Femto, Plasma Surface Technology) at 100 W for 3 min and then moved to a vacuum desiccator (Bel-Art F420220000, Thermo Fisher Scientific, 08-594-16B) for chemical vapor deposition (CVD). PDMS-silane (1,3-dichlorotetramethylsiloxane; Gelest, SID3372.0) (25 μL ×2 per treatment) was vaporized under pumping for 3 min and then condensed onto glass substrate under vacuum at room temperature for 40 min. The PDMS-grafted surface was thoroughly rinsed with ethanol (anhydrous, 99.5%), deionized (DI) water, and then dried with nitrogen for use. Step #2) Fabrication of PDMS stamps. Photomasks were designed in Adobe Illustrator and then sent to a service (Fineline Imaging) for printing. Standard photolithography was applied to make a master that contains all the microchannel features. Details about photolithography can be found in our previous publication^30^. Last, PDMS stamps were made by pouring a degassed (about 20 min using a vacuum desiccator) silicone precursor and curing agent mix (SYLGARD 184, Silicone Elastomer Kit, Dow, 04019862) in 10:1 mass ratio onto the master and cured on a hotplate at 80 °C for 4 h. The PDMS stamps were stripped off with tweezers and punched with through holes (Miltex Biopsy Punch with Plunger, Ted Pella, 15110) at the inlet and outlet of a microchannel for the following O_2_ plasma surface patterning. Step #3) O_2_ plasma surface patterning. The PDMS silane-grafted chamber coverglass was masked by a punched PDMS stamp and then treated with O_2_ plasma at 100 W for 1 min. After surface patterning, the PDMS stamp was removed by tweezers and stored in a clean container (e.g., Petri dish) for reuse. Step #4) Under-oil microchannels without ECM coating - The chemically patterned chambered coverglass from Step #3 was overlaid with 1 mL silicone oil (<100 cSt, e.g., 5 cSt or 20 cSt) for each well on the 2-well chambered coverglass. The target media was distributed onto the microchannels by under-oil sweep. Briefly, get 20 μL of the target media in a 1-200 μL large orifice pipet tip (02-707-134, Thermo Fisher Scientific) and then drag the hanging microdrop at the end of the tip through the patterned surface. Media was spontaneously distributed onto the O_2_ plasma-treated areas only. For double-oil overlay [i.e., silicone oil (20 cSt) + fluorinated oil (Fluorinert FC-40)], we first prepared the under-oil microchannels under silicone oil (20 cSt) with the target media by under-oil sweep. Next, we added 1 mL Fluorinert FC-40 (1.85 g/mL at 25 °C) for each well on the 2-well chambered coverglass directly into the silicone oil (20 cSt, 0.95 g/mL at 25 °C). Due to the high density of the fluorinated oil and immiscibility with silicone oil, the fluorinated oil spontaneously replaces silicone oil in a well and pushes it to the top layer. At last, the silicone oil was removed using pipet at the four corners in a well. Step #5) Under-oil microchannels with ECM coating - Like Step#4, the under-oil microchannels with ECM coating were prepared by sweeping a hanging microdrop of the ECM solution under oil (silicone oil, 5 cSt). The ECM solution is collagen I (Rat Tail, about 10 mg/mL in the original bottle) 1:1 dilution in 2% N-2-hydroxyethylpiperazine-N-2-ethane sulfonic acid (HEPES) 2× PBS and then 1:1 dilution in RPMI (basal media only). The final pH of the collagen solution was adjusted to 7.4 by 0.5 M sodium hydroxide (NaOH) endotoxin-free aqueous solution (0.5 μL of 0.5 M NaOH per 100 μL collagen solution). After under-oil sweep, the device was kept at room temperature (∼21 °C) for 1 h for polymerization. The ECM layer effectively retains hydration of the microchannels with single-oil overlay (Fig. 1c). After the ECM layer is fully polymerized, the device is ready for sample loading. Step #6) Sample loading before imaging. The device with oil overlay was set up under a microscope. We first registered the xy and z positions of the microchannels with perfect focal plane (PFS) function. Next, we loaded the blood or cell stock using a pipet under oil for 1 μL/spot (Fig. 1b) throughout all the microchannels. At last, we loaded the chemoattractant solution or pathogen inoculum at the other end of each microchannel for 1 μL/spot. After sample loading and update of the xy and z positions, the device is ready for imaging or timelapse.

### Whole blood collection

All blood samples were drawn according to Institutional Review Boards (IRB) approved protocols per the Declaration of Helsinki at the University of Wisconsin-Madison in the Microtechnology, Medicine, and Biology (MMB) Lab (IRB# 2020-1623, healthy donors) and in the Lang Lab (IRB# 2014-1214, cancer patients). Informed consent was obtained from all subjects in the study. Whole blood was collected with standard Vacutainer tubes (EDTA tube, BD, 366643; ACD tube, BD, 364606) (Supplementary Fig. 9a). Finger prick blood (90 μL) was collected using a pipet and following the standard finger prick lancet (Microtainer, contact-activated, BD, 366594) protocol into a 0.6 mL sterile Eppendorf tube pre-loaded with 10 μL of ACD-A solution (Sigma Aldrich, C3821-50 mL) (Supplementary Fig. 9b). All the whole blood samples were stored in a standard CO_2_ incubator (Thermo Fisher Scientific, HERACELL VIOS 160i) at 37 °C before use.

### Neutrophil isolation

Neutrophils were isolated from whole blood using magnetic bead-based negative selection. Two negative selection kits - MACSxpress Whole Blood Neutrophil Isolation Kit, human (Miltenyi Biotec, 130-104-434) and EasySep Direct Human Neutrophil Isolation Kit (STEMCELL, 19666) - were compared. In MACSxpress isolation, RBCs were further depleted from the neutrophil pellet using BD Pharm Lyse buffer, according to the manufacturer’s instruction. In EasySep isolation, the neutrophil pellet was directly resuspended in the target media for further experiments. Due to the relatively low non-specific activation of neutrophils from isolation (Supplementary Fig. 4c), EasySep was used throughout the μ-Blood project. RPMI (basal media only) was used as the isolation buffer. Cell count of each isolation was obtained using a hemocytometer (LW Scientific) (Supplementary Fig. 6c).

### Autologous plasma separation

Autologous plasma was separated from the same whole blood sample along with neutrophil and PBMC isolations. To get PFP (i.e., platelet-free plasma), the whole blood tube was spun at 1000 *g*/10 min (Eppendorf, Centrifuge 5702, brake on) first. The supernatant was transferred to several 1.5 mL sterile Eppendorf tubes at 1 mL/tube. The supernatant tubes were then spun at 10000 *g*/10 min (Eppendorf, Centrifuge 5424, brake on). The supernatant (i.e., PFP) in the Eppendorf tubes was aliquoted and stored at a -80 °C freezer (Thermo Fisher Scientific, Forma 900 Series) for future use. The aliquots needed in an experiment were thawed at room temperature (∼21 °C) with only one freeze-thaw use.

### MAGPIX neutrophil-related cytokine analysis

Multiplexed protein secretion analysis was performed with plasma samples (PFP, see *Autologous plasma separation* above) and conditioned media, using the MAGPIX Luminex Xmap system (Luminex) with a custom-built bead panel (Thermo Fisher Scientific, Human ProcartaPlex Mix&Match 18-plex, PPX-18-MXZTFFM), per the manufacturers’ protocol. Data of conditioned media are pooled from 3 replicates. Samples were frozen at -80 °C from the time of collection until the assay was performed. Secreted protein levels were displayed as z-scores in a heat map generated by GraphPad Prism 10.0.1.

### Microscopic imaging

Bright-field, fluorescence images and videos were recorded on a Nikon Ti Eclipse inverted epifluorescence microscope (Nikon Instruments). The chambered coverglass with oil overlay was kept at 37 °C, 21% O_2_, 5% CO_2_, and 95% RH in an onstage incubator (Bold Line, Okolab) during imaging or timelapse. After imaging on the microscope, the μ-Blood devices were moved to and kept in a standard CO_2_ incubator (37 °C, 18.6% O_2_, 5% CO_2_, and 95% RH) before the characterization for the next time point.

### Area fraction analysis

The fluorescence images of Sytox and Annexin V (4× magnification) were batch processed and analyzed using a customized Java code (Github - https://github.com/jcai0791/Neutrophil-Project) calling functions from Fiji ImageJ (Supplementary Fig. 8a). The workflow includes: 1) Open Fiji ImageJ. 2) Analyze → Set Measurements… (select “Area fraction”). 3) Open a set of the original .nd2 images with Sytox and Annexin V channels (Image → Type → 16-bit). 4) Image → Duplicate… (Separate the Sytox and Annexin V channels by making a duplicate). 5) Process → Subtract Background… (Rolling ball radius: 50 pixels). 6) Image → Adjust → Threshold… (pick a Threshold Method, Default for the Sytox channels, or MaxEntropy for Annexin V channels, select “Dark background”). 7) Analyze → Measure. 8) File → Save As → Jpeg… (use “default file name+Threshold Method-Sytox or Annexin V”). 9) Save Results (File → Save as… “file name.csv”). The data points in the .csv datasheet files were organized in the order of time points with conditions under each time point from the group of Sytox or Annexin V channels.

|  | 0 h | 1 h | ... |
| --- | --- | --- | --- |
| Condition a | / | / | / |
| Condition b | / | / | / |
| ... | / | / | / |

10) To plot the data points, we transposed the datasheet in the order of conditions with time points under each condition from the group of Sytox or Annexin V channels.

|  | Condition a | Condition b | ... |
| --- | --- | --- | --- |
| 0 h | / | / | / |
| 1 h | / | / | / |
| ... | / | / | / |

11) The sorted data were plotted in GraphPad Prism 10.0.01.

### Clumping analysis

The fluorescence images of Sytox (4× magnification) were batch processed and analyzed using a customized Java code (Github - https://github.com/jcai0791/Neutrophil-Project) calling functions from Fiji ImageJ (Supplementary Fig. 8b-e). The workflow includes: Each image was first cropped, then split into a grid of GRIDSIZE pixels. The mean fluorescence intensity of each grid square was measured and then plotted in GraphPad Prism 10.0.1.

### Cell counting and speed tracking analysis

Manual counting was performed in Fiji ImageJ with the timelapse videos recorded on the Nikon microscope by counting each individual neutrophil that passes into the channel. For speed tracking, images in a timelapse were transferred into Fiji ImageJ, where cell tracking was performed manually using the Fiji plugin “MTrackJ”. Once “MTrackJ” is selected, the “add” option allows tracking of each individual migrating cell. Once the manual tracking is done for each migrating cell, measurements can be taken automatically. Measurements were created through the “MTrackJ” measurements option.

### Statistical analysis

Raw data was directly used in statistical analysis with no data excluded. Data was averaged from at least 3 replicates (unless otherwise stated) and present as mean ± standard deviation (S.D.) if applicable. The statistical significance was specified in the figure captions. All statistical analyses were performed using GraphPad Prism 10.0.1.

## Data availability

The data that support the findings of this study are available from the corresponding authors upon reasonable request.

## Acknowledgements

This project was performed in the MMB Lab at the University of Wisconsin-Madison with the support from Dr. David J. Beebe and Dr. Anna Huttenlocher. We thank Dr. Jennifer Schehr (The Lang Lab at the University of Wisconsin-Madison) for the discussion on the influence of anticoagulants on neutrophil migration and collection of cancer blood samples, Mrs. Alice Golubiewski, Mr. Ravi Chandra Yada, and Dr. Cristina Sanchez de Diego (The MMB Lab at the University of Wisconsin-Madison) for arranging and collecting the healthy blood samples, Mr. Terry Juang (The MMB Lab at the University of Wisconsin-Madison) for demonstrating the finger prick blood collection workflow, Mrs. Sue McCrone (The Rose Lab at the University of Wisconsin-Madison) for providing the standard *S. aureus* inoculum. This work was supported by NIH R01 CA247479, NIH R01 AI154940, NIH R01 EB010039, NIH P30CA014520, NIH U24AI152177, and NCI R01 CA085862.

## Author contributions

C.L. conceived the project and designed the research. C.L. developed the μ-Blood assay platform and performed data collection, analysis, and visualization with the assistance from N.W.H., M.M., M.A.F., Z.A., J.K., and E.A.W.. T.J. and C.L. prepared the 3D model of the under-oil microchannel with neutrophil migration. C.L. supervised the project. C.L. wrote the manuscript and all authors revised it.

## Competing interests

The authors declare no competing financial interests.

## Notes

### Competing Interest Statement

The authors have declared no competing interest.

### Author Declarations

All blood samples were drawn according to Institutional Review Boards (IRB) approved protocols per the Declaration of Helsinki at the University of Wisconsin-Madison in the Microtechnology, Medicine, and Biology (MMB) Lab (IRB# 2020-1623, healthy donors) and in the Lang Lab (IRB# 2014-1214, cancer patients). Informed consent was obtained from all subjects in the study.

### Summary of Updates

Updated authorship and funding information. Reorganized figures.

