## Supplementary material for "Microliter whole blood neutrophil assay preserving physiological lifespan and functional heterogeneity": SI

**This PDF file includes:**

SI Sections I to VI
Supplementary Figures 1 to 10
Supplementary Table 1
Legends for Supplementary Movies 1 to 5
SI References

**SI Section I - The challenges of achieving micrometer-scale open channels for open-fluid single-cell trapping**

One unique challenge in open microfluidics through nearly the last two decades was the highly limited lateral resolution, e.g., channel width limited to millimeter scale and above (Supplementary Fig. 1). It is easy to print a smooth and consistent liquid filament (or channel) on a chemically homogenous (i.e., untextured, unpatterned) substrate in air (similar to using a marker on whiteboard) but channels with channel width less than one millimeter dry up quickly in a few minutes or seconds even in a humidified environment (Supplementary Fig. 1-I), which along with the vulnerability to airborne contaminants limits the use of single-liquid-phase channels in micro total analysis system (μTAS) and biomedical assays. To minimize evaporation and contamination during liquid printing, printing under oil seems to be a straightforward solution, however, Plateau-Rayleigh instability (i.e., a liquid filament tends to spontaneously break up to micro droplets)[1] makes printing a smooth and consistent liquid channel on a chemically homogeneous substrate, especially with an oil overlay and a large aspect ratio of the channel, very challenging to nearly impossible[2] (Supplementary Fig. 1-II). Using a chemically patterned (i.e., heterogeneous) surface with wettability contrast (e.g., superhydrophobic-superhydrophilic) can effectively suppress the effect of Plateau-Rayleigh instability during liquid printing[3]. However, the superhydrophobic-superhydrophilic patterned surfaces are still limited by its apparent liquid repellency [i.e., liquid repellency enabled by surface structures with Young's contact angle, $\theta$ < 180$^{o}$ and thus local fouling and sample loss especially when bio-liquids (i.e., sticky and easy fouling) are used][4] and compromised optical access and mechanical durability[5] (Supplementary Fig. 1-III). By contrast, chemically patterned, untextured surfaces with double exclusive liquid repellency (Double-ELR) provide absolute, inherent liquid repellency (with Young's contact angle, $\theta$ = 180$^{o}$) with uncompromised optical access and mechanical durability[2] (Supplementary Fig. 1-IV). Under-oil microchannels with micrometer-scale lateral resolution (defined by pre-patterning, see Methods) can be rapidly and robustly prepared by simply sweeping a hanging drop (at any size) across a Double-ELR surface by hand-held pipet[2]. There might be thoughts of using liquid wicking rather than printing to make open microchannels because there are many techniques available to fabricate dry channels on various substrates with micrometer- to nanometer-scale lateral resolution, e.g., photolithography, soft lithography, physical/chemical vapor deposition, and e-beam lithography (Supplementary Fig. 1-V, VI). The challenge of liquid wicking is the limited wicking length both in air (primarily due to evaporation) and under oil (primarily due to the oil displacement energy barrier)[2].

Overall, to the best of our knowledge Double-ELR-enabled under-oil sweep distribution[2] is the only reported method so far that allows lossless, rapid, and high-precision preparation of micrometer-scale open microfluidic channels with various bio-liquids (e.g., standard culture media, collagen solution, serum/plasma, cellular and microbial suspensions) and advanced fluid controls (including open-fluid single-cell trapping, high flow rate range, and on-demand reversible open-fluid valves). These features in ELR-empowered UOMS support various *in vitro* cell culture/interrogation microenvironments and assays with low adoption/implementation barriers.

Here, we test a series of the Double-ELR under-oil microchannels in experiments for open-fluid single-cell trapping of blood cells and bacteria (Supplementary Fig. 2). The microchannels consist of a straight channel connecting two circular spots, with the dimensions written in the format of, e.g., 2-2-*L*0.5-*W*0.1 (w or w/o ECM), for 2 mm in diameter of the spots, 0.5 mm in channel length (*L*) and 0.1 mm in channel width (*W*). The channel width of interest in this study is ≤100 μm with a channel length ≤500 μm. Whole blood [white blood cells, 7 to 30 μm in diameter; RBCs, 7.5 to 8.7 μm in diameter and 1.7 to 2.2 μm in thickness; platelets, 3 to 4 μm in diameter] and bacteria [*S. aureus*, 0.5 to 1.5 μm in diameter] are used and tested regarding the immune cell functional assays. Without ECM coating, the channel height of 2-2-*L*0.5-*W*0.1 is calculated for 2.3 μm (Supplementary Fig. 2b, inset). In this case, RBCs can get passively pumped through the microchannel as a monolayer of cells with the disc in parallel to the substrate surface, evidenced by the assembly, disassembly, and re-assembly of RBC rouleaux (i.e., stacks of RBCs) before, in, and out of the microchannel (Supplementary Fig. 2c, top left). The following loading of *S. aureus* (GFP-labeled) at the opposite spot showed a confined entrance plume due to the occupation of channel space by RBCs (Supplementary Fig. 2c, top right). The other microchannels with a smaller channel width of 30-40 μm all showed complete blood cell trapping (Supplementary Fig. 2c, bottom left) due to the reduced channel height to <0.3 μm (Supplementary Fig. 2b, inset). It is worth noting that if the channel length is smaller than 2× of the length of entrance plume ($l_e$), RBCs can still get passively pushed through the microchannel (Supplementary Fig. 2c, bottom right). In comparison, all the microchannels tested with ECM coating trapped both blood cells and bacteria (Supplementary Fig. 2d) on the seeding spots due to the much thinner media layer compared to the ECM-free conditions [Supplementary Fig. 2b, inset; media layer thickness of 2-2-*L*0.5-*W*0.1 (w/o ECM) is 2.3 μm; media layer thickness of 2-2-*L*0.5-*W*0.1 (w ECM) is 2.3 μm - 0.7 μm = 1.6 μm].

**SI Section II - Non-specific activation of isolated neutrophils**

Here we demonstrate the non-specific activation of isolated neutrophils from a pilot study (Supplementary Fig. 4). Human primary neutrophils were isolated directly from a whole blood sample with negative selection (Supplementary Fig. 4a, Methods). Right after isolation, rather than interrogated with operator-defined stimuli (e.g., cytokines, cells, pathogens, and drugs), the cells were directly cultured on a 384-well plate (30K cells per well) in standard neutrophil culture media [RPMI + 10% FBS + 1% Pen/Strep, 20 μL per well] in a standard $CO_2$ incubator (37 ºC, 18.6% $O_2$, 5% $CO_2$, 95% RH). Surprisingly, we observed that more than 50% of the cells were activated non-specifically in just a few hours (<4 h) from an isolation since the initiation of cell culture (Supplementary Fig. 4b). The activation process can be easily identified by the change of cell morphology from round ball (i.e., live stand-by cells, 10 μm in diameter) to flat disc (i.e., dead cells, 15-20 μm in diameter) under bright field (Fig. 2d). Further visualized with Sytox (a membrane impermeable DNA stain) and Hoechst (a membrane permeable DNA stain), neutrophils in all the possible cell stages can be identified including live (i.e., "healthy") cells, early apoptosis, secondary necrosis, suicidal NETosis, and vital NETosis (Supplementary Fig. 4b, Fig. 2e). A further investigation on the non-specific activation was performed with a group of 7 donors consisting of healthy donors (self-reported) and cancer patients (Supplementary Fig. 4c). The results revealed a highly divergent non-specific activation of isolated neutrophils varying from <5% to >50% in 2 to 4 h in standard 2D neutrophil monoculture (Supplementary Fig. 4d).

Further, a systematic review of the complete workflow of *in vitro* study of human neutrophils led us to a large parameter space with >15 variables including whole blood history, isolation method, and culture conditions that may affect/alter neutrophil kinetics and activation. In two parallel projects, we quantitatively evaluated and compared the effect of these variables on non-specific activation of neutrophils. The conclusion is that from *in vivo* to *ex vivo* neutrophils can get easily, randomly, and non-specifically activated (i.e., without operator-defined stimuli) due to the altered environmental factors. Such inconsistencies associated with neutrophil isolation sparked the idea of developing a new immune cell functional assay method that directly uses unprocessed whole blood as both the assay input and through the assays for readout collection. Keeping neutrophils in their whole blood with the autologous signaling molecules, constituent cells, and the physiological level of oxygen (e.g., 5% $O_2$ in venous blood) allows removal of all the operation steps in neutrophil isolation and minimized disturbance on those extremely sensitive cells, leading to improved extraction of donor-specific information and efficiency, consistency, and throughput of the assays.

**Section III - Under-oil evaporation and double-oil overlay**

Published under-oil evaporation test[6]: The retention of water was tested with 1 μL ELR water droplets (diameter = 1.24 mm, surface area = 4.83 $mm^2$) with a single-oil overlay of silicone oil (5 cSt, 5 mm in thickness) in an incubator (37 °C, 40% humidified). The droplets lasted a week on average before completely evaporating. The average evaporation rate of water in this condition was estimated around 1 nL/h/$mm^2$. The media volume in the microchannel 2-2-$L$0.5-$W$0.1 (w/o ECM) (surface area = 0.5 mm × 0.1 mm = 0.05 $mm^2$) is known as 257.62 pL right after sweep (Supplementary Table. 1). If we assume no volume compensation to the microchannel from the two spots via lateral flow, the media volume of 0.26 nL lasts about 0.26 nL/(0.05 $mm^2$ × 1 nL/h/$mm^2$) ≈ 5 h. If we consider that the evaporation rate of the 1 μL ELR water droplets in the beginning of the experiment should be much higher than the average evaporation rate (due to the largest surface area), the media volume in the channel will last much shorter than 5 h.

In μ-Blood evaporation test (Supplementary Fig. 5), we used the microchannel 2-2-$L$0.5-$W$0.1 (w/o ECM) with a 1.5 mm overlay of silicone oil (5 cSt) (i.e., 1 mL oil in a well of a standard Lab Tek II 2-well chambered coverglass) (Fig. 1a). The microchannels were prepared with PFP by under-oil sweep and then loaded with whole blood and housed in an onstage incubator (37 °C, 21% $O_2$, 5% $CO_2$, 95% RH). As discussed and shown in Supplementary Fig. 2c, RBCs get passively pushed into this microchannel from whole blood loading onto the spot. The volume of PFP in the microchannel occluded by RBCs is 1/2 × 0.26 nL × 90% (water content of plasma) ≈ 0.12 nL. The microchannel with RBCs dried up in about 20 min from the start of imaging, which gives an average evaporation rate of 0.12 nL/0.05 $mm^2$/0.33 h ≈ 7 nL/h/$mm^2$ in μ-Blood with a single-oil overlay and no ECM coating. The microchannels with ECM coating stay hydrated for at least 12 h tested in another project (not included in this manuscript). By contrast, the microchannels, even without ECM coating stay hydrated for more than 6 days with the double-oil overlay [i.e., silicone oil (20 cSt) + fluorinated oil (Fluorinert FC-40)] due to the significantly lowered moisture permeability through fluorinated oil (Supplementary Fig. 5b).

**SI Section IV - Donor heterogeneity and N1 versus N2 polarization**

Our immune system is highly adaptive to the environment; therefore, person-to-person heterogeneity of neutrophils is expected. In a parallel project, we collected whole blood samples from 36 donors including healthy donors (self-reported) and cancer patients of different etiologies (Supplementary Fig. 6a). To quantify the functional heterogeneity of neutrophils in whole blood, we analyzed 18 neutrophil-related cytokines (pro-inflammatory, anti-inflammatory, regulatory, and chemoattractants)[7] in the plasma of each whole blood sample (Supplementary Fig. 6b). Healthy donors showed more heterogeneous secretion of cytokines compared to cancer patients. Cytokine concentrations from neutrophils in cancer patients were lower compared to the healthy donors, which is indicative of tumor-directed/altered neutrophil function[8]. In comparison, the neutrophil-related cytokine expression profiles in the healthy donors fell along a more disparate spectrum, which could be related to the donor's health condition during blood collection[9].

We further compared the neutrophil counts from the donor pool and the influence of isolation method and whole blood storage conditions (Supplementary Fig. 6c). The average neutrophil count of healthy donors and cancer patients in this project were 1.5 ± 1.1 versus 2.0 ± 1.1 million/mL in whole blood. It is worth noting that the average yield (i.e., isolated neutrophils/neutrophils in whole blood ×100%) of the EasySep Direct Kit is about 50% (information from STEMCELL). It is known that cancer is typically associated with an elevated neutrophil-to-lymphocyte ratio and neutrophil count[10]. Comparison between MACSxpress and EasySep isolation kits showed no statistical difference in neutrophil counts. Short-time whole blood storage (<8 h) did not lower neutrophil count. However, long-time whole blood storage (>24 h) led to decreased neutrophil counts across different donors.

Next, we looked across 60 neutrophil isolations from 36 donors by comparing non-specific activation of neutrophils in standard 2D neutrophil monoculture (RPMI + 10% FBS +1% Pen/Strep) on tissue culture-treated polystyrene 384-well plate in a standard $CO_2$ incubator.

Neutrophils from healthy donors showed significantly higher sensitivity to the environmental factors from *in vivo* to *ex vivo* as evidenced by the high non-specific activation (Supplementary Fig. 7a). Phenotypically, neutrophils from the healthy donors showed a higher frequency (~20% versus ~0%) in forming clumps compared to neutrophils from cancer patients (Supplementary Fig. 7b,c).

Recent studies on neutrophil diversity and plasticity have identified N1 and N2 subtypes of neutrophils which possess pro-inflammatory and anti-inflammatory phenotypes[11], respectively. There is also significant skewing of neutrophils to N1 over N2 or vice versa in autoimmune disease and certain cancer types[12]. Here we hypothesize that neutrophils from patients with cancer would have a more anti-inflammatory N2 subtype. Furthermore, neutrophils from cancer patients are thought to have decreased sensitivity to their environment/stimuli and could therefore be less prone to non-specific activation. We hypothesize that the desensitization could be attributed to the neutrophil polarization (i.e., N1 versus N2) state.

To investigate these hypotheses, isolated neutrophils from a donor were partitioned into three groups - no treatment, N1 polarization, and N2 polarization. We adopted the N1 and N2 polarization protocol published by M. Ohms *et. al*[13] and compared the clumping phenotype and non-specific activation between the negative control group (no treatment) and the two polarized groups (Supplementary Fig. 7d,e). The N1-polarized group showed a higher incidence of clumping relative to the negative control group. By contrast, the N2-polarized group showed no significant difference on clumping compared to the negative control group, but interestingly, had decreased non-specific activation.

These pilot studies on donor heterogeneity and neutrophil polarization extended to the μ-Blood study using unprocessed whole blood, aiming for improved extraction of donor-specific information and assay consistency.

**SI Section V - Influence of anticoagulants on neutrophil recruitment**

Anticoagulants are commonly used to prevent blood clotting after blood draw by interfering with the normal blood clotting process. While anticoagulants have been used to prevent or treat thromboembolic events, there is evidence that they can affect other physiological processes, including cell migration. Anticoagulants affect cell migration by altering the function of certain signaling molecules or enzymes that are involved in the migration process. For example, heparin and warfarin inhibit the activity of proteases, which are involved in the breakdown of the ECM and the release of signaling molecules[14]. Ethylenediaminetetraacetic acid (EDTA)[15] - a divalent cation (such as $Mg^{2+}$, $Ca^{2+}$, $Mn^{2+}$, $Fe^{2+}$, and $Zn^{2+}$ ions) chelator - disrupts the divalent-cation-dependent integrin binding by removing the cations. Similarly, anticoagulant citrate dextrose (ACD)[16] acts as an anticoagulant by the action of the citrate anion chelating $Ca^{2+}$. Here we test the influence of EDTA and ACD at their blood collection concentration - EDTA at 1.8 mg/mL of blood; ACD-A solution (22.0 mg/mL trisodium citrate, 8.0 mg/mL citric acid, and 24.5 mg/mL dextrose) at 1:10 dilution with blood - on neutrophil recruitment in μ-Blood.

The results showed that EDTA at the standard blood collection concentration (1.8 mg/mL of blood) strongly inhibited neutrophil migration (Supplementary Fig. 9, Supplementary Movie 1), which is attributed to its chelation and depletion of a broad range of bivalent cations. In comparison, ACD-A at the working concentration (1:10 dilution with blood) still allowed neutrophil migration (Supplementary Fig. 9). This reflects the narrow, $Ca^{2+}$-specific chelation of ACD. In these tests, the blood samples were collected via venipuncture (Supplementary Fig. 9a). While venipuncture is the standard in most blood assays, it requires a specialist and a designated area (e.g., in hospitals, clinics, or point-of-care settings) to perform blood draw. In addition, venipuncture is an invasive method that can be limited by donor conditions (e.g., donors with difficult veins, newborns) and comes with the risk of infection if not performed properly.

To expand the adaptability of μ-Blood, we explored the feasibility of using finger prick (Supplementary Fig. 9b) that is more flexible on operator-donor conditions, locations, and less invasive and safer. While the volume that can be collected typically from one finger pick (a few tens of microliters) is much lower than venipuncture (normally several milliliters), it is not a limitation to μ-Blood because μ-Blood only requires 1 μL or below per testing unit (e.g., a microchannel). A few tens of microliters are already more than enough to cover all the microchannels on a device (e.g., 24 channels per Lab Tek-II chambered coverglass) (Fig. 1a). To work with finger prick blood, the clotting time of whole blood (typically 2-8 min) needs to be considered, which defines the blood sample loading window and throughput. For manual sample loading on a μ-Blood device by pipetting, it takes about 10-15 s per loading therefore about 1 min per 4 microchannels or 6 min per device (with 24 channels) (Fig. 1a) with an average training in experimental operations. If using a robotic fluid dispenser[17], the sample loading can be much faster and therefore supports higher throughput. Our results showed that neutrophil recruitment was not affected from finger prick blood without anticoagulants (Supplementary Fig. 9c, Supplementary Movie 1) at least in 2-4 h of neutrophil migration. It is worth noting that the finger prick blood on a device should have fully coagulated without anticoagulant in several hours, but neutrophils still respond to and migrate toward a chemoattractant gradient. The compatibility of μ-Blood with anticoagulant-free finger prick blood allows more flexible blood collection and operator-donor conditions, and immune cell functional assays where anticoagulants are a major concern. Anticoagulants (e.g., ACD solution) can be added to finger prick blood like venipuncture blood. With anticoagulants, the limitation on the sample loading window due to blood clotting is gone.

**SI Section VI - Influence of channel dimensions on sample loading and neutrophil recruitment**

ELR-empowered UOMS allows highly flexible surface pattern selection[2]. In this work, we focus on the simplest channel geometry - a straight channel connecting two circular spots for sample loading (e.g., whole blood/blood cells, chemoattractant, pathogen) (Fig. 1). The spot size, channel length, and channel width can be designed/adjusted independently (Supplementary Fig. 2a). The channel height [ECM (channel height@"II-Equilibrium") + media (channel height@"IV-Equilibrium" minus channel height@"II-Equilibrium")] with ECM coating or media only (channel height@"IV-Equilibrium") without ECM coating] of a given design is determined by Laplace pressure equilibrium between the channel and the spots after sample loading (Supplementary Fig. 2b).

In μ-Blood, the under-oil microchannels were prepared by "under-oil sweep" - dragging a hanging drop of the target media (e.g., culture media, collagen solution, plasma) across the patterned Double-ELR surface. Right after sweep, all the liquid takes a nearly constant height-to-width ($H/W$) ratio of 1/13, leading to a Laplace pressure differential between the channel ($\Delta P_{channel} = \gamma_{oil\text{-}media}/R_{channel\text{-}width}$) and the spots ($\Delta P_{spot} = 2\gamma_{oil\text{-}media}/R_{spot}$) (Supplementary Fig. 2a, I-Right after sweep) [2]. In this work, the channel width (100 μm or below) is much smaller than the spot diameter (2 mm) so liquid gets pumped out of the channel to the spots until Laplace pressure equilibrium is reached (Supplementary Fig. 2a, II-Equilibrium). Samples are then loaded onto the spots, which breaks the Laplace pressure equilibrium (Supplementary Fig. 2a, III-Add 1 μL/spot of media) and pumps liquid back to the channel until the system reaches a new Laplace pressure equilibrium (Supplementary Fig. 2a, IV-Equilibrium).

In our previous work[2], we quantified the mass transport through an under-oil microchannel using a fluorescent probe molecule - 2-NBDG (fluorescence-labeled glucose, M.W. 342.26 g/mol). Briefly, we prepared the channels (2-2-$L$/$W$, see below) by under-oil sweep and then loaded 0.5 μL of 2-NBDG solution (2 mM in media) to the inlet spot. The flow rate ($Q$, pL/min) was achieved by monitoring/analyzing the fluorescence intensity change over time on the outlet spot as follows:

2-2-$L$0.137-$W$0.058 (w/o ECM), $Q$ = 225 pL/min

2-2-*L*0.135-*W*0.058(w ECM), *Q* = 81 pL/min

2-2-*L*0.413-*W*0.035 (w/o ECM), *Q* = 18 pL/min
2-2-*L*0.422-*W*0.042 (w ECM), *Q* = 13 pL/min

Due to the ultra-low volume in the microchannels used in μ-Blood - e.g., the volume in the channel right after sweep: 2-2-*L*0.5-*W*0.1 (257.6 pL) and 2-2-*L*0.26-*W*0.04 (13.9 pL) (Supplementary Table. 1) - the system reaches Laplace pressure equilibrium quickly in about 1 min after under-oil sweep or sample loading.

On the μ-Blood microchannels (2-2-*L*0.5-*W*0.1 and 2-2-*L*0.26-*W*0.04), we load whole blood (1 μL) first to all the microchannels on a device (e.g., about 6 min to finish sample loading on 24 channels per chambered coverglass device) (Fig. 1a) and then load the chemoattractant solution (1 μL) with Hoechst (1:200 dilution for nucleus staining/visualization during timelapse) to the other spot. The volume variation between the first microchannel and the last microchannel from sample loading on a device (e.g., with *Q* = 225 pL/min, the volume change is 1.35 nL in 6 min, or *Q* = 18 pL/min, the volume change is 0.1 nL in 6 min) is negligible compared to 1 μL (i.e., the sample loading volume) thanks to the micrometer-scale channel dimensions (Supplementary Fig. 2b) and the ultrasmall flow rate (listed above), which is an important enabling factor that allows a manageable operation window for consistent sample loading.

The gradient establishment of a given chemoattractant (i.e., molecular weight, concentration) at the working temperature (e.g., 37 ºC in this study) in a microchannel is determined by the channel dimensions and the ECM layer in the channel. Another factor that affects the under-oil microchannel environment and thus neutrophil migration is under-oil media loss via evaporation (see Section III in SI for detailed discussion). The measured evaporation rate in μ-Blood with a single-oil overlay and no ECM coating is about 7 $nL/h/mm^2$. For ultrasmall sample/fluid volumes, the media loss via evaporation under oil cannot be ignored. To minimize under-oil evaporation and retain the hydration of the microchannels, several strategies have been developed in our research including i) introducing ECM (e.g., collagen I) coating to the spots and the microchannels[18]. The ECM hydrogel layer effectively locks water molecules in the 3D matrix and facilitates media compensation from the spots via enhanced capillary wicking (compared to ECM-free channels); ii) using silicone oil with higher viscosity (e.g., 1000 cSt, 10,000 cSt); and iii) using a double-oil (e.g., silicone oil + fluorinated oil) overlay (Supplementary Fig. 5b). Oil with high viscosity and fluorinated oil both effectively and significantly reduce media loss via evaporation under oil. At last, with a given established gradient of chemoattractant and minimized media loss via evaporation, the channel width defines the maximum number of neutrophils at a migration front ($N_{\text{front}}$) in a microchannel, e.g., $N_{\text{front}}$ = 3-8 on 2-2-*L*0.5-*W*0.1 (Supplementary Movie 2-1, Supplementary Movie 2-3), and $N_{\text{front}}$ = 1-3 on 2-2-*L*0.26-*W*0.04 (Supplementary Movie 2-2).

The experimental results of the influence of channel dimensions and chemoattractant (fMLP) concentrations on neutrophil recruitment are summarized in Supplementary Fig. 10. In the microchannels 2-2-*L*0.5-*W*0.1 (w ECM), 2-2-*L*0.26-*W*0.04 (w ECM), and 2-2-*L*0.07-*W*0.04 (w/o ECM), wider and/or shorter channels recruited more neutrophils with a similar migration speed at around average 0.3 μm/s (Supplementary Fig. 10a-c). Similarly, in the microchannels 2-2-*L*0.16-*W*0.04 (w/o ECM) and 2-2-*L*0.26-*W*0.04 (w/o ECM), shorter channels also recruited more neutrophils with channel length showing no significant influence on migration speed (Supplementary Fig. 10d-f). Increased fMLP concentration (100 nM → 200 nM → 300 nM) led to a comparable or reduced migration speed from average 0.2 μm/s to average 0.15 μm/s. In the microchannels 2-2-*L*0.5-*W*0.1 (w/o ECM), 2-2-*L*0.26-*W*0.04 (w/o ECM), 2-2-*L*0.16-*W*0.04 (w/o ECM), and 2-2-*L*0.07-*W*0.04 (w/o ECM), wider and/or shorter channels recruited more neutrophils with the wider *W*0.1 microchannels showing a significantly higher migration speed (average 0.25 μm/s) compared to the *W*0.04 microchannels (average 0.15 μm/s) (Supplementary

Fig. 10g-i). Increased fMLP concentration (0.1 μM → 1 μM → 10 μM) led to a reduced migration speed in *W*0.1 microchannels from average 0.25 μm/s to average 0.2 μm/s, and in the *W*0.04 microchanenls from average 0.15 μm/s to average <0.1 μm/s. Again, channel length showed no significant influence on migration speed. The selection of a specific channel dimension in an experiment is clarified in the main text.

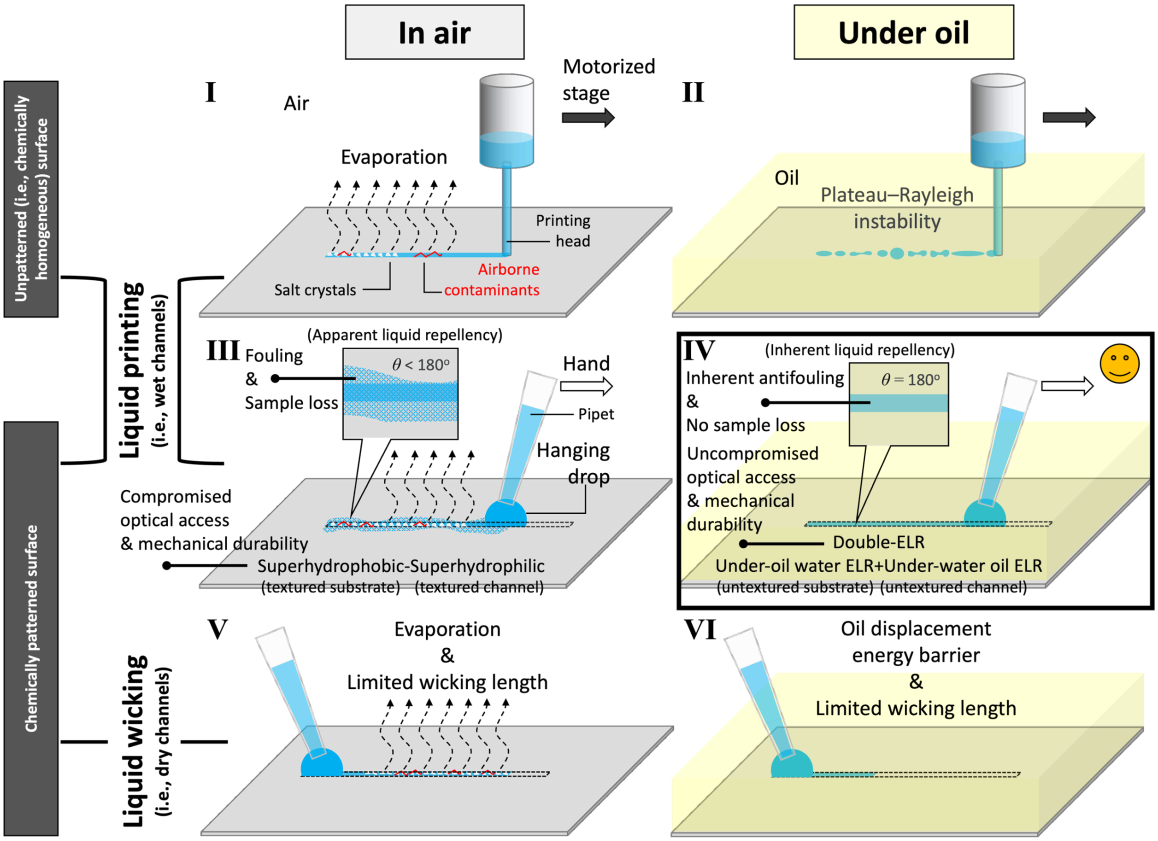


**Supplementary Fig. 1.** Why is making sub-1 mm (wide) open-fluid channels challenging? The left column shows the preparation of open-fluid channels in air - (from top to bottom) liquid printing on chemically homogenous surface (I), liquid printing on chemically patterned (i.e., heterogeneous) surface (III), and liquid wicking on chemically patterned surface (V). The right column shows the preparation of open-fluid channels under oil (II, IV, and VI) corresponding to each method listed in the left column.

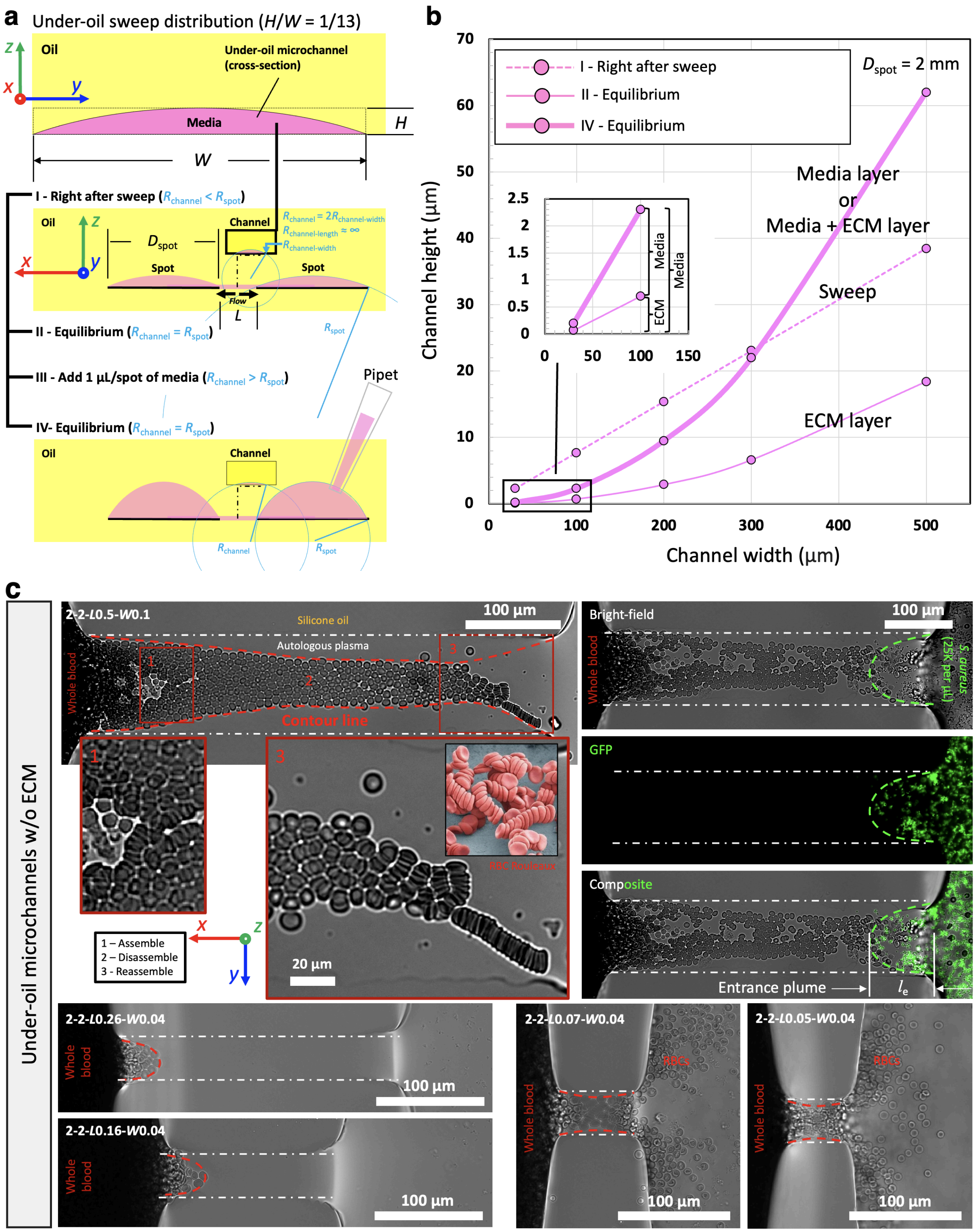
a
Under-oil sweep distribution (H/W = 1/13)
Oil
Under-oil microchannel (cross-section)
Media
H
W
I - Right after sweep (Rchannel < Rspot)
II - Equilibrium (Rchannel = Rspot)
III - Add 1 µL/spot of media (Rchannel > Rspot)
IV- Equilibrium (Rchannel = Rspot)
Channel
Spot
Flow
L
Pipet
b
Dspot = 2 mm
I - Right after sweep
II - Equilibrium
IV - Equilibrium
Media layer or Media + ECM layer
Sweep
ECM layer
Channel height (µm)
Channel width (µm)
c
Under-oil microchannels w/o ECM
2-2-L0.5-W0.1
Silicone oil
Autologous plasma
Whole blood
Contour line
1 – Assemble
2 – Disassemble
3 – Reassemble
RBC Rouleaux
20 µm
100 µm
Bright-field
GFP
Composite
S. aureus (25K per µL)
Entrance plume
le
2-2-L0.26-W0.04
2-2-L0.16-W0.04
2-2-L0.07-W0.04
2-2-L0.05-W0.04
RBCs

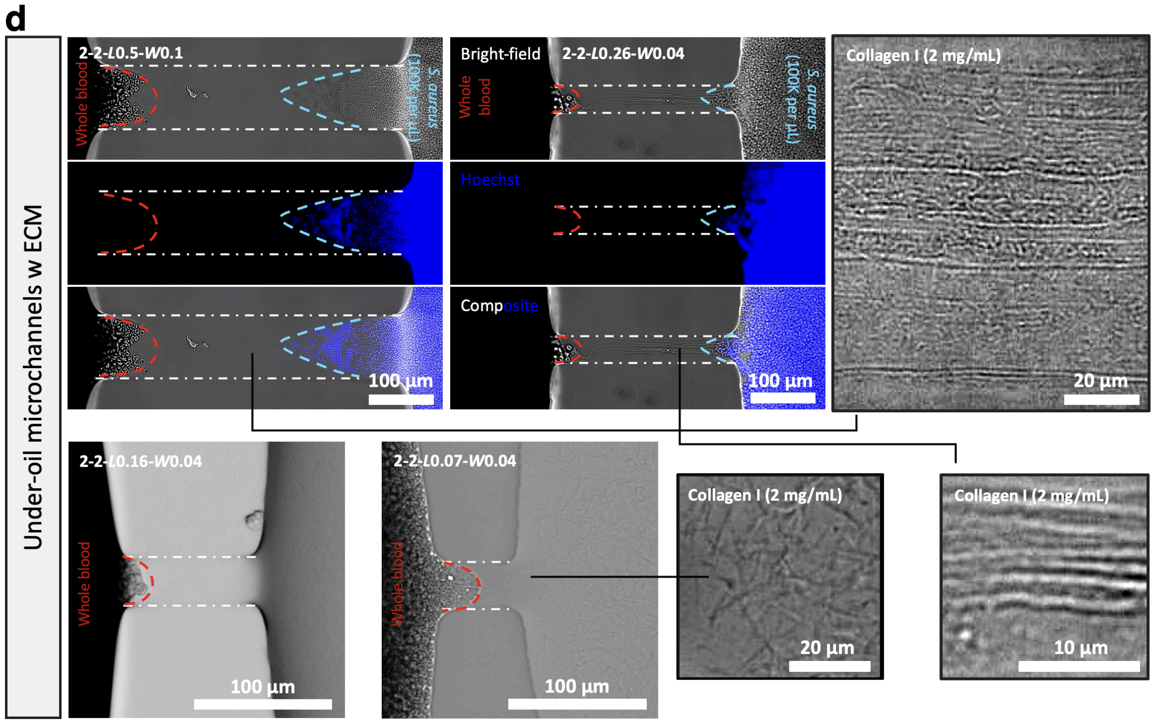


**Supplementary Fig. 2.** Laplace pressure equilibrium analysis of the under-oil microchannel and experimental results of open-fluid single-cell trapping. a) Schematics that show the four stages of fluid dynamics including I-Right after sweep, II-(Laplace pressure) equilibrium, III-Add 1 µL/spot of media, and IV-(Laplace pressure) equilibrium. Right after sweep (I), all the liquid layers take a nearly constant height-to-width ($H/W$) ratio [2] of 1/13. Driven by the Laplace pressure differential between the microchannel [$\Delta P_{microchannel} = \gamma_{oil\text{-}media}/R_{channel\text{-}length} + \gamma_{oil\text{-}media}/R_{channel\text{-}width} = \gamma_{oil\text{-}media}/R_{channel\text{-}width}$ (with $R_{channel\text{-}length} \rightarrow \infty$), where $\Delta P$ is Laplace pressure, $\gamma_{oil\text{-}media}$ is the interfacial tension at the oil-meida interface, $R$ is the radius of curvature] and the spots ($\Delta P_{spot} = 2\gamma_{oil\text{-}media}/R_{spot}$), the fluid volume in the microchannel (Supplementary Table. 1) gets pumped out until the system reaches equilibrium (II). Adding an extra volume (1 µL in this work) to a spot (III) breaks the equilibrium, pumping some volume back to the microchannel until the system reaches a new equilibrium (IV). b) Theoretically calculated channel height as a function of channel width for spot diameter of 2 mm. (Inset) A zoomed-in graph of the channel width ≤100 µm. For the microchannels with ECM coating, the thickness of the ECM layer is described by "II-Equilibrium" and the thickness of the media layer on top of the ECM layer can be known as the differential between "IV-Equilibrium" and "II-Equilibrium". For the microchannels without ECM coating, the thickness of the media layer is described by "IV-Equilibrium". c), d) Open-fluid single-cell trapping results in the under-oil microchannels without ECM (c) and with ECM coating (d). The channel dimensions are the measured values on a microscope (Supplementary Table. 1, footnote). The channel oil-media boundaries on the substrate are highlighted by the white dash-dotted lines. The entrance plumes (i.e., the surface area occupied by the cells in a microchannel, Fig. 1c) are marked out by the contour lines (colored dashed lines: red - whole blood; green - GFP-labeled *S. aureus*, blue - *S. aureus* stained with Hoechst). When the channel length is smaller than 2× of the entrance plume length ($l_e$), RBCs get passively pushed through the channel.

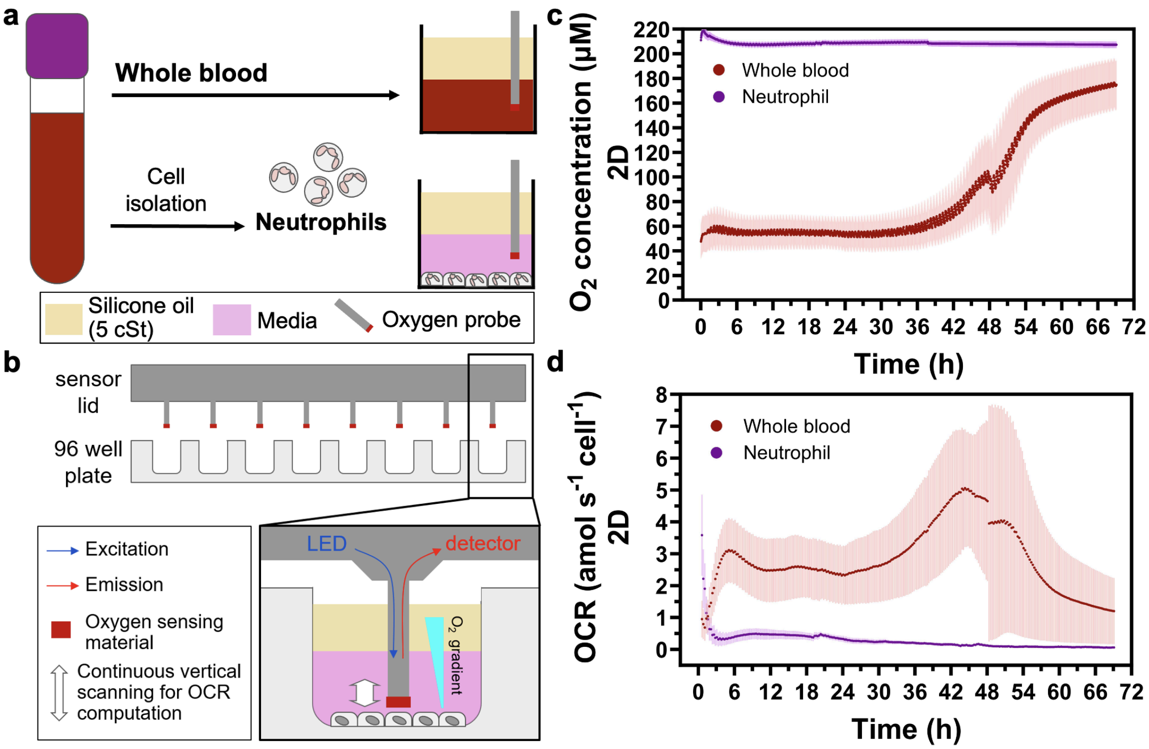


**Supplementary Fig. 3.** Comparison of oxygen kinetics between whole blood and isolated neutrophils in UOMS cell culture. a) Schematic shows the 2D under-oil culture microenvironment on a standard 96-well plate. Whole blood was used directly unprocessed for 120 µL/well. Isolated neutrophils were seeded at $10^6$ cells/cm$^2$ in a volume of 120 µL/well of culture media (RPMI + 10% FBS + 1% Pen/Strep). All the experimental groups were overlaid with 100 µL/well of silicone oil (5 cSt). b) Schematic shows the Resipher system (an automated oxygen optical sensor unit) and the measurement of c) oxygen concentration and d) oxygen consumption rate (OCR). Results are expressed as mean ± S.D. from 3 replicates.

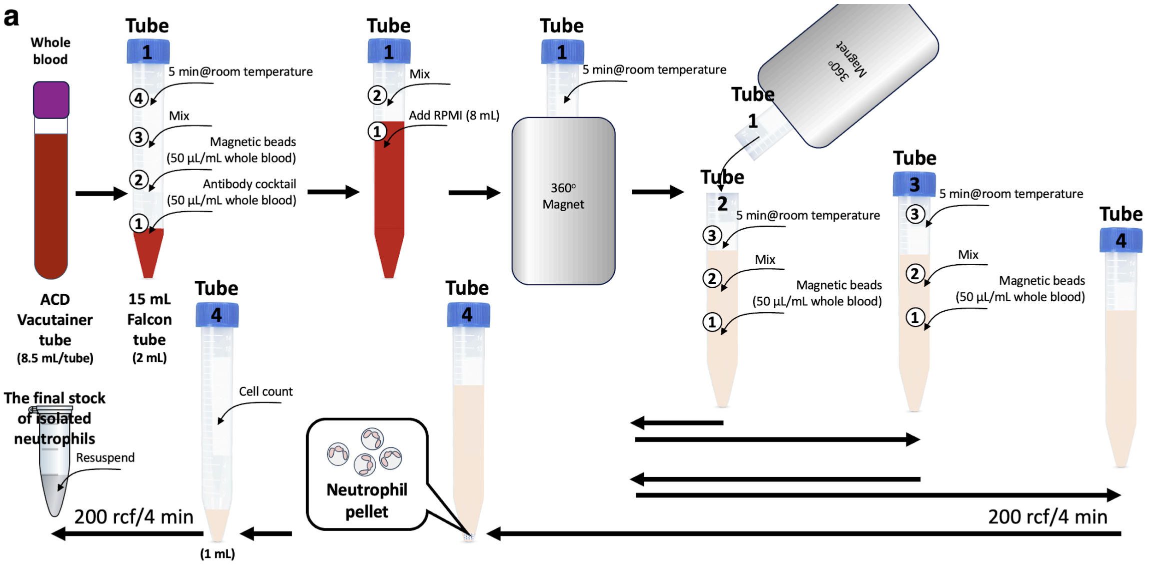

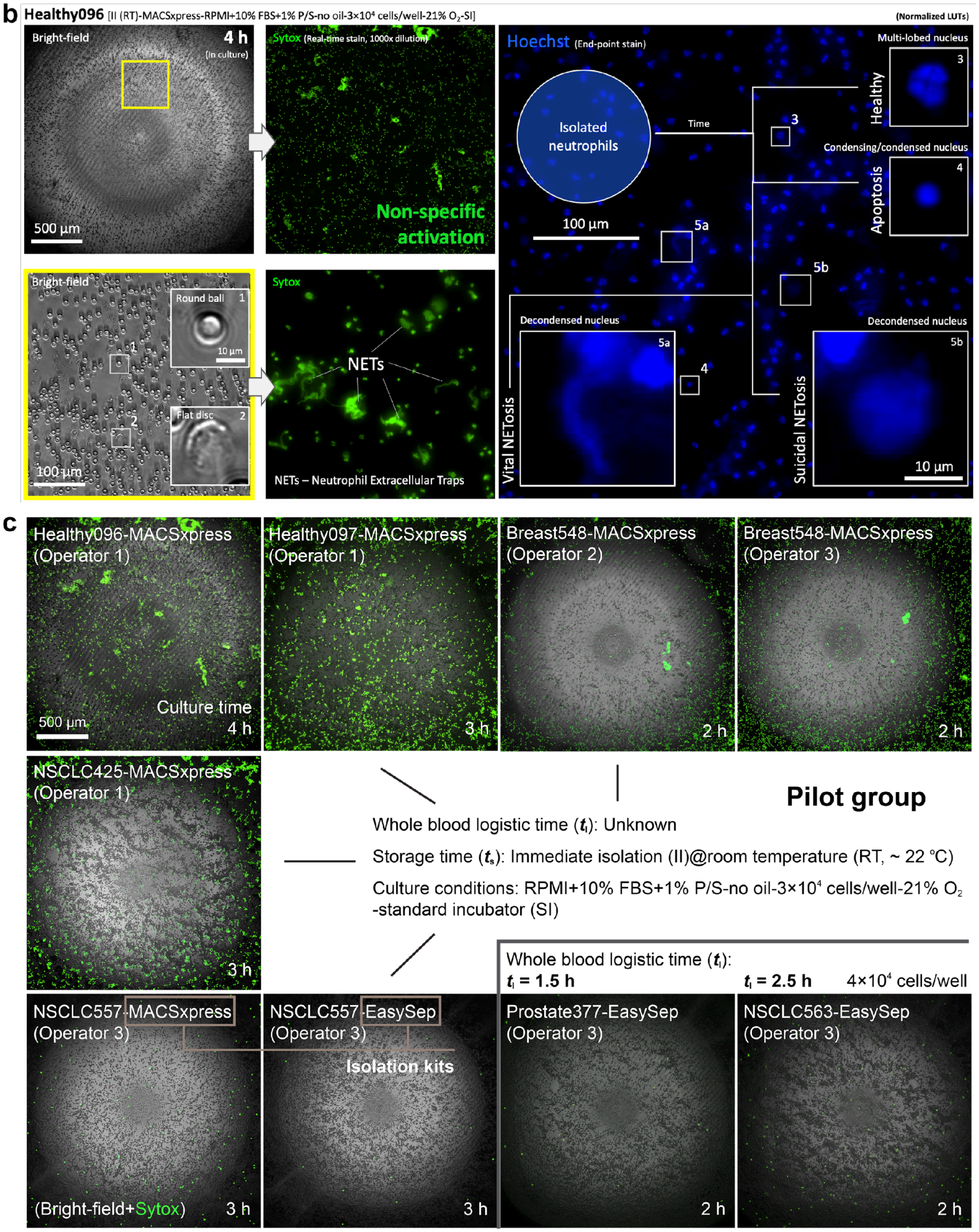
b
Healthy096 [II (RT)-MACSxpress-RPMI+10% FBS+1% P/S-no oil-3×10⁴ cells/well-21% O₂-SI]
(Normalized LUTs)
Bright-field
4 h
(In culture)
500 µm
Sytox (Real-time stain, 1000x dilution)
Non-specific activation
Bright-field
Round ball 1
10 µm
Flat disc 2
100 µm
Sytox
NETs
NETs – Neutrophil Extracellular Traps
Hoechst (End-point stain)
Isolated neutrophils
Time
100 µm
Multi-lobed nucleus
Healthy
Condensing/condensed nucleus
Apoptosis
Decondensed nucleus
Vital NETosis
Decondensed nucleus
Suicidal NETosis
10 µm
c
Healthy096-MACSxpress (Operator 1)
Culture time 4 h
500 µm
Healthy097-MACSxpress (Operator 1)
3 h
Breast548-MACSxpress (Operator 2)
2 h
Breast548-MACSxpress (Operator 3)
2 h
NSCLC425-MACSxpress (Operator 1)
3 h
Pilot group
Whole blood logistic time (tₗ): Unknown
Storage time (tₛ): Immediate isolation (II)@room temperature (RT, ~ 22 °C)
Culture conditions: RPMI+10% FBS+1% P/S-no oil-3×10⁴ cells/well-21% O₂ -standard incubator (SI)
Whole blood logistic time (tₗ):
tₗ = 1.5 h
tₗ = 2.5 h
4×10⁴ cells/well
NSCLC557-MACSxpress (Operator 3)
(Bright-field+Sytox)
3 h
NSCLC557-EasySep (Operator 3)
Isolation kits
3 h
Prostate377-EasySep (Operator 3)
2 h
NSCLC563-EasySep (Operator 3)
2 h

d

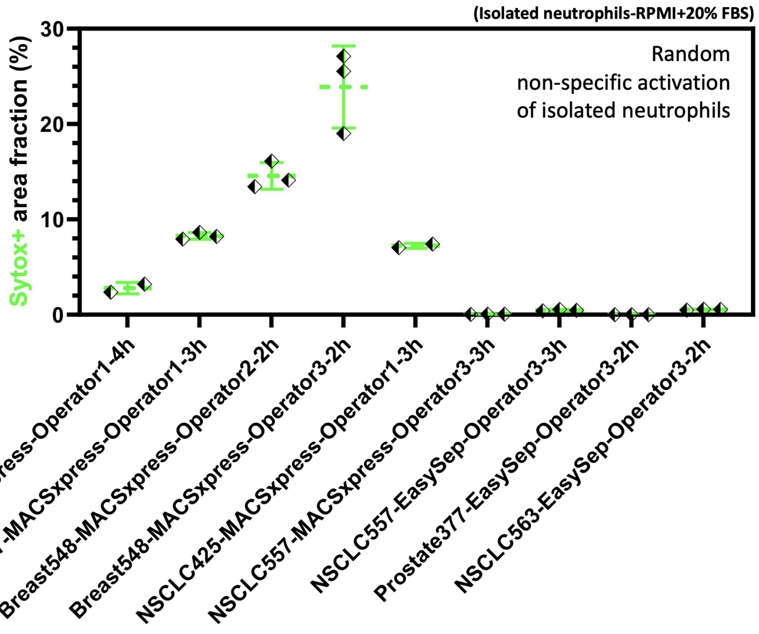


**Supplementary Fig. 4.** The results of non-specific activation of isolated neutrophils in a pilot study. a) The workflow of typical negative selection-based direct neutrophil isolation from whole blood (Methods), taking 20+ steps and >1 h per isolation from whole blood to the final stock of isolated neutrophils. b) The first observation in the pilot study of high non-specific activation (>50%) from a healthy donor. c) The variation of non-specific activation (from 7 donors consisting of healthy donors and cancer patients) ranging from <5% to >50% in 2 to 4 h of standard 2D neutrophil monoculture after neutrophil isolation. d) Quantified results of non-specific activation of isolated neutrophils (i.e., area fraction of Sytox+ cells, Supplementary Fig. 8a, Methods) in (c). Error bars are mean ± S.D..

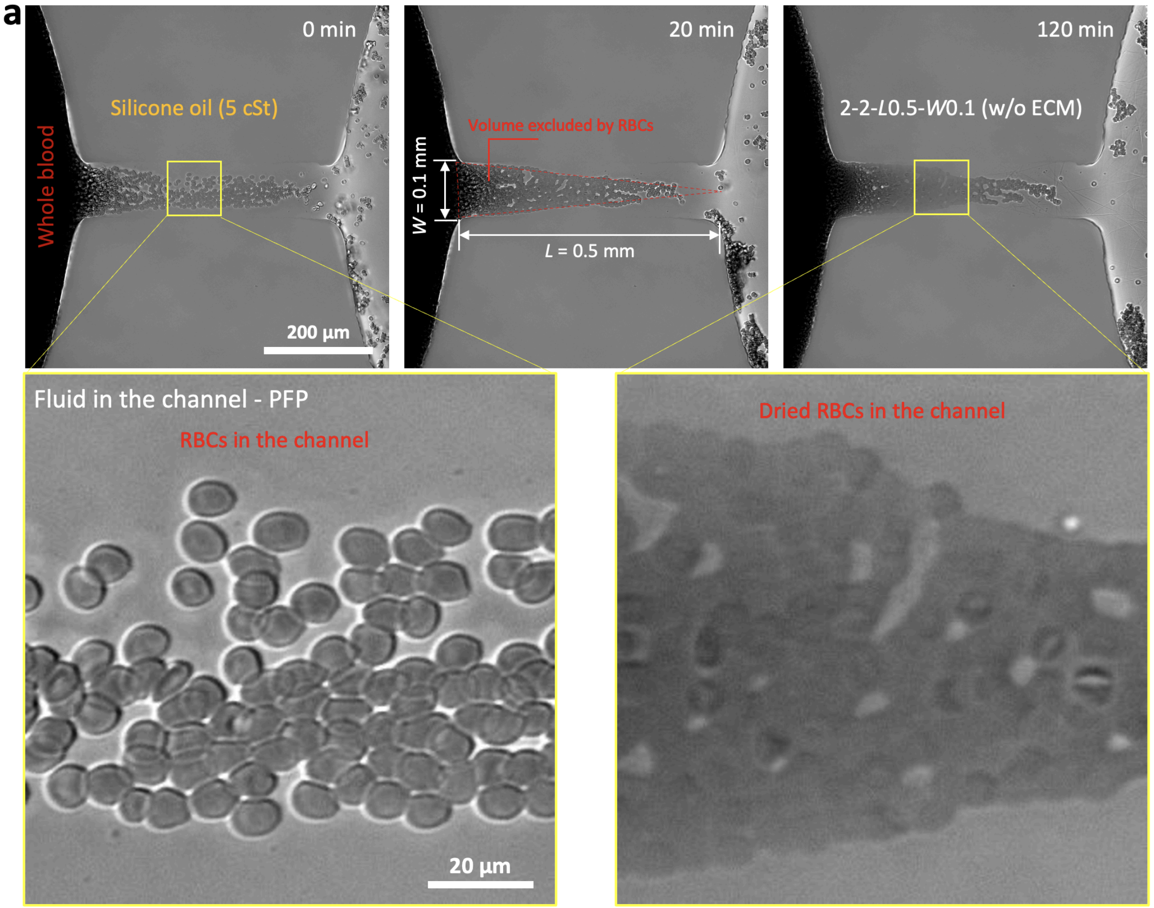
a
0 min
20 min
120 min
Silicone oil (5 cSt)
Whole blood
Volume excluded by RBCs
W = 0.1 mm
L = 0.5 mm
2-2-L0.5-W0.1 (w/o ECM)
200 µm
Fluid in the channel - PFP
RBCs in the channel
20 µm
Dried RBCs in the channel

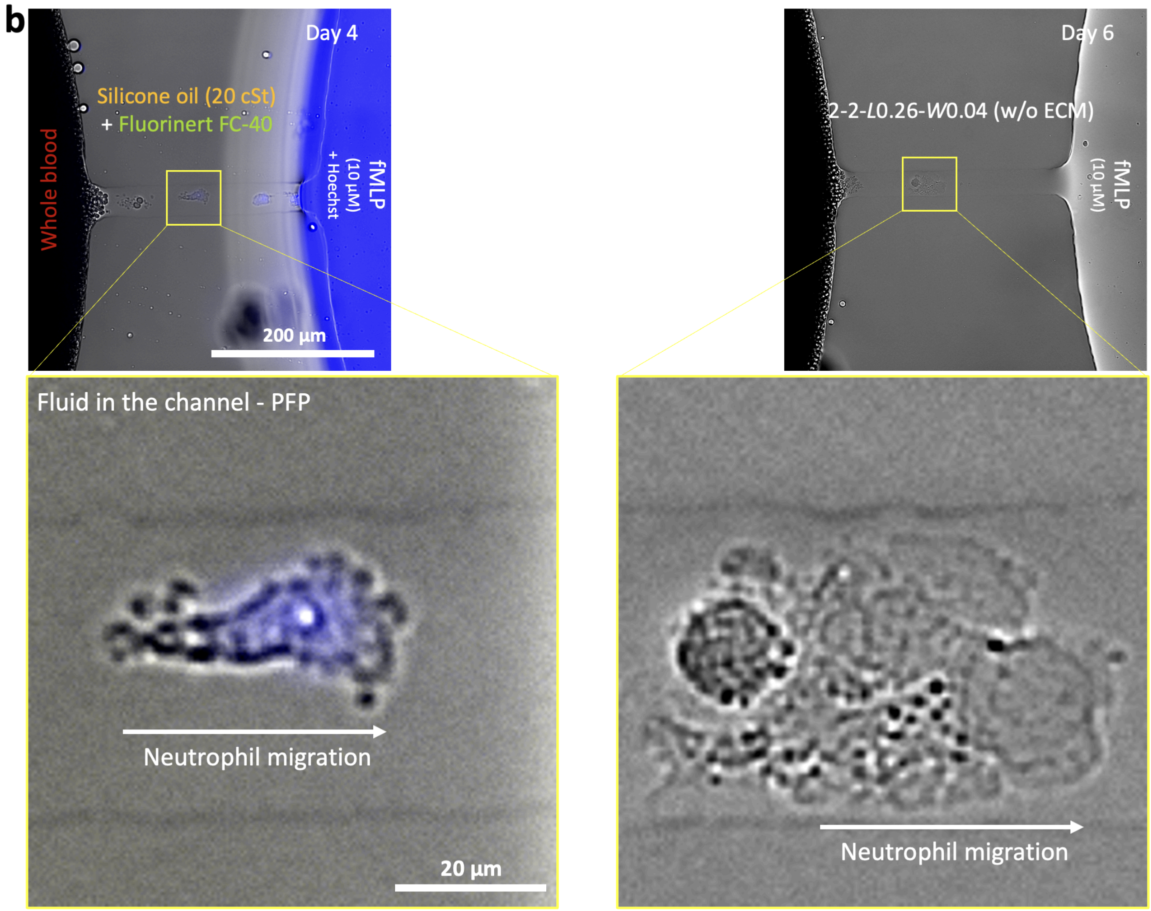


**Supplementary Fig. 5.** Media loss via evaporation under oil and the comparison between single-oil and double-oil overlay in µ-Blood. a) Media loss via evaporation in the under-oil microchannels 2-2-*L*0.5-*W*0.1 (w/o ECM) with single-oil overlay (2 h). b) Media retention in the under-oil microchannels 2-2-*L*0.26-*W*0.04 (w/o ECM) with double-oil overlay (6 days) (Supplementary Movie 5).

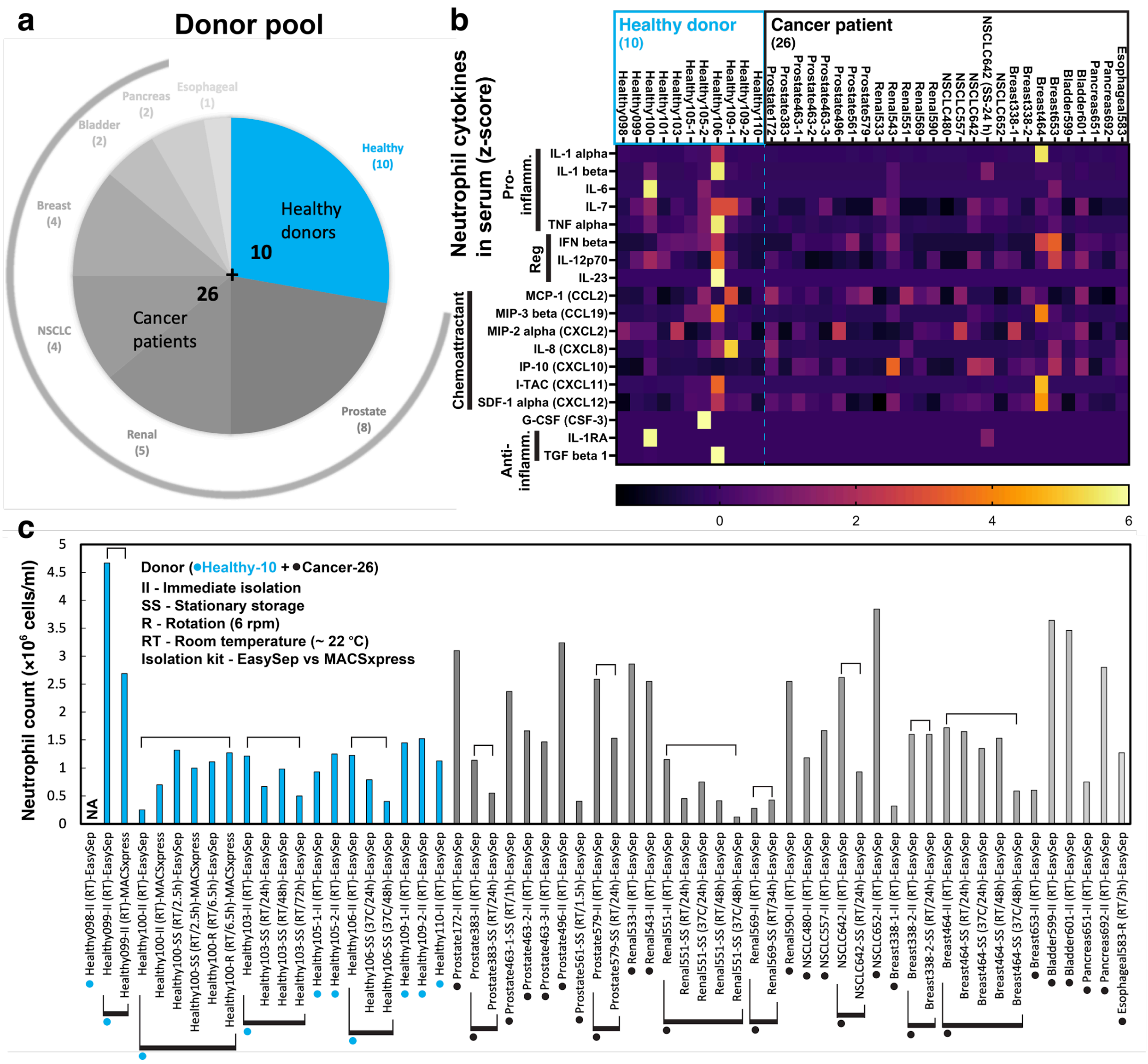


**Supplementary Fig. 6.** Statistics of the donor pool in the donor heterogeneity study. a) The donor pool consists of 10 healthy donors and 26 cancer patients. b) Heat map (z-score) of 18-plex neutrophil-related cytokines in autologous plasma (Methods). c) Neutrophil counts from 60 isolations show the variation among donors, between the two isolation kits (MACSxpress versus EasySep, Methods) used for negative selection-based neutrophil isolation from whole blood, and the influence of storage method (stationary storage versus rotation), time, and temperature of whole blood.

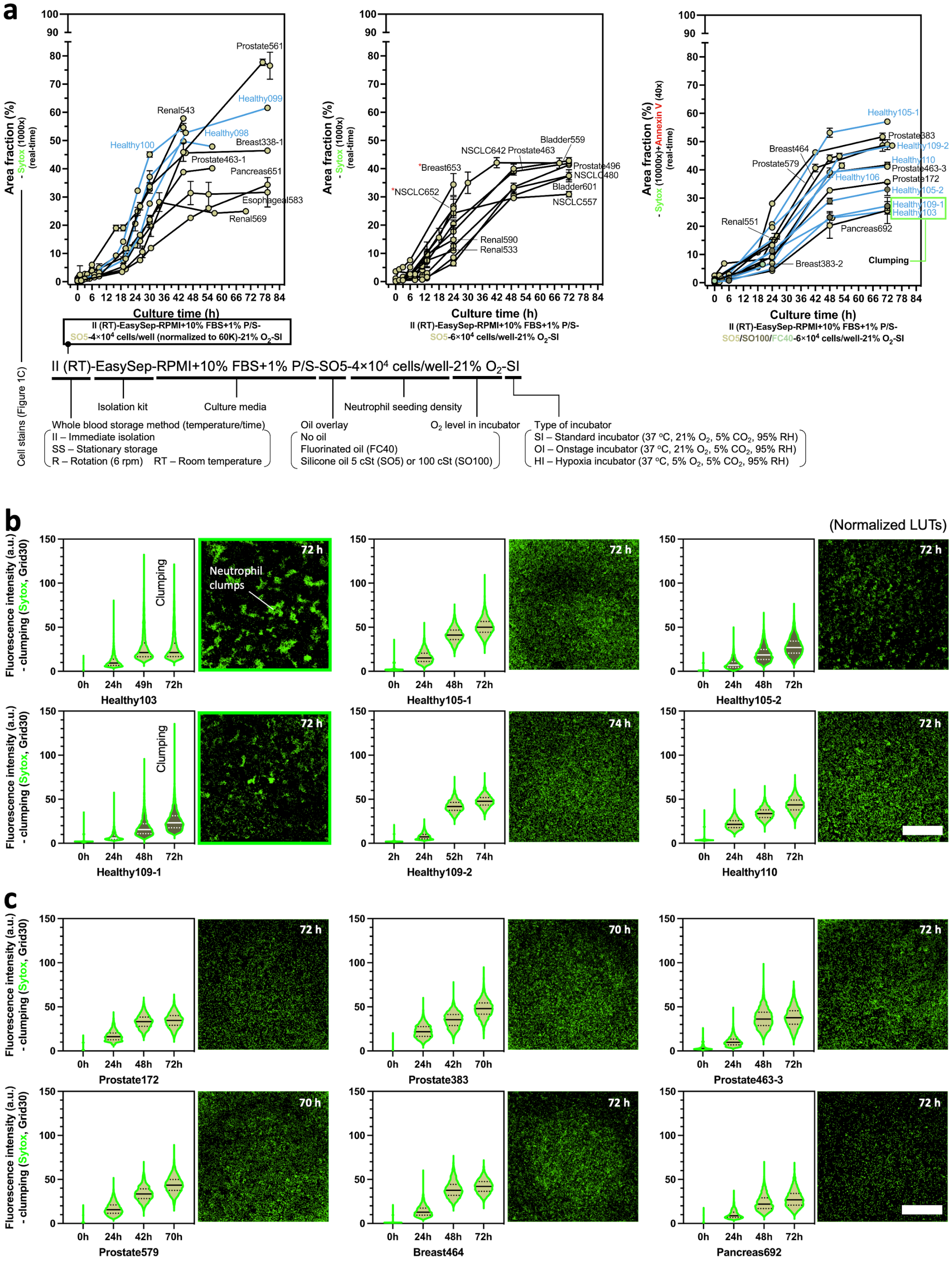
a
Area fraction (%) - Sytox (1000x) (real-time)
Culture time (h)
Prostate561
Healthy099
Renal543
Healthy098
Healthy100
Breast338-1
Prostate463-1
Pancreas651
Esophageal583
Renal569
II (RT)-EasySep-RPMI+10% FBS+1% P/S-SO5-4×10⁴ cells/well (normalized to 60K)-21% O₂-SI
Bladder559
NSCLC642
Prostate463
Breast653
NSCLC652
Prostate496
NSCLC480
Bladder601
NSCLC557
Renal590
Renal533
II (RT)-EasySep-RPMI+10% FBS+1% P/S-SO5-6×10⁴ cells/well-21% O₂-SI
Area fraction (%) - Sytox (10000x)+Annexin V (40x) (real-time)
Healthy105-1
Prostate383
Healthy109-2
Breast464
Prostate579
Healthy110
Prostate463-3
Healthy106
Prostate172
Healthy105-2
Healthy109-1
Healthy103
Renal551
Pancreas692
Breast383-2
Clumping
II (RT)-EasySep-RPMI+10% FBS+1% P/S-SO5/SO100/FC40-6×10⁴ cells/well-21% O₂-SI
II (RT)-EasySep-RPMI+10% FBS+1% P/S-SO5-4×10⁴ cells/well-21% O₂-SI
Isolation kit
Culture media
Neutrophil seeding density
Whole blood storage method (temperature/time)
II – Immediate isolation
SS – Stationary storage
R – Rotation (6 rpm) RT – Room temperature
Oil overlay
No oil
Fluorinated oil (FC40)
Silicone oil 5 cSt (SO5) or 100 cSt (SO100)
O₂ level in incubator
Type of incubator
SI – Standard incubator (37 °C, 21% O₂, 5% CO₂, 95% RH)
OI – Onstage incubator (37 °C, 21% O₂, 5% CO₂, 95% RH)
HI – Hypoxia incubator (37 °C, 5% O₂, 5% CO₂, 95% RH)
Cell stains (Figure 1C)
b
(Normalized LUTs)
Fluorescence intensity (a.u.) - clumping (Sytox, Grid30)
Clumping
Neutrophil clumps
Healthy103
Healthy105-1
Healthy105-2
Healthy109-1
Healthy109-2
Healthy110
c
Prostate172
Prostate383
Prostate463-3
Prostate579
Breast464
Pancreas692

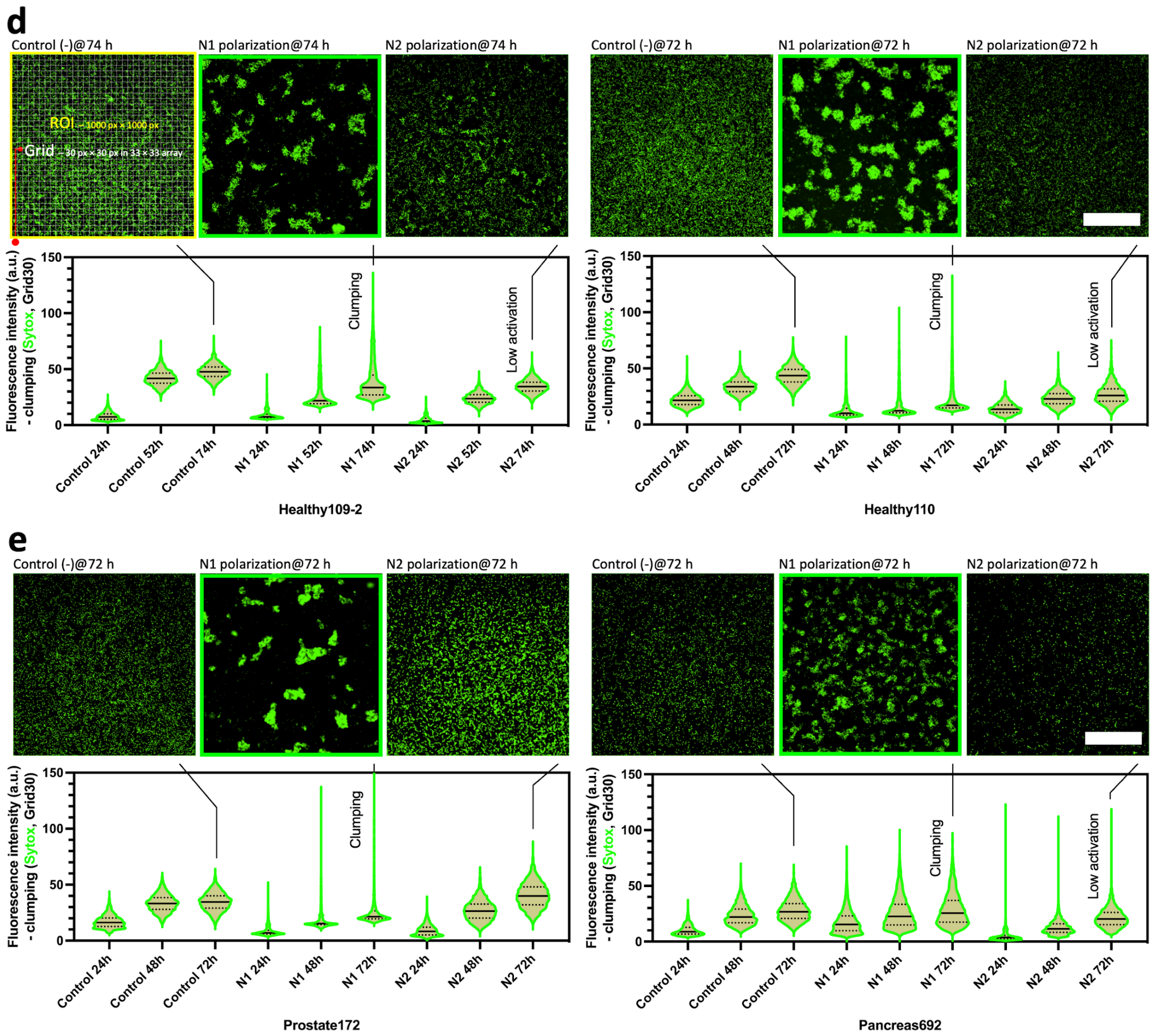


**Supplementary Fig. 7.** Donor heterogeneity (healthy donors versus cancer patients) identified and quantified by non-specific activation of neutrophils. a) Area fraction (Supplementary Fig. 8a) results from the Sytox (real-time staining) channel. b) and c) Clumping analysis (Supplementary Fig. 8b-e) of the representative healthy donor group and cancer patient group compared to d) and e) N1 versus N2 polarization tests. Scale bar, 500 µm. Error bars are mean ± S.D. from ≥3 replicates.

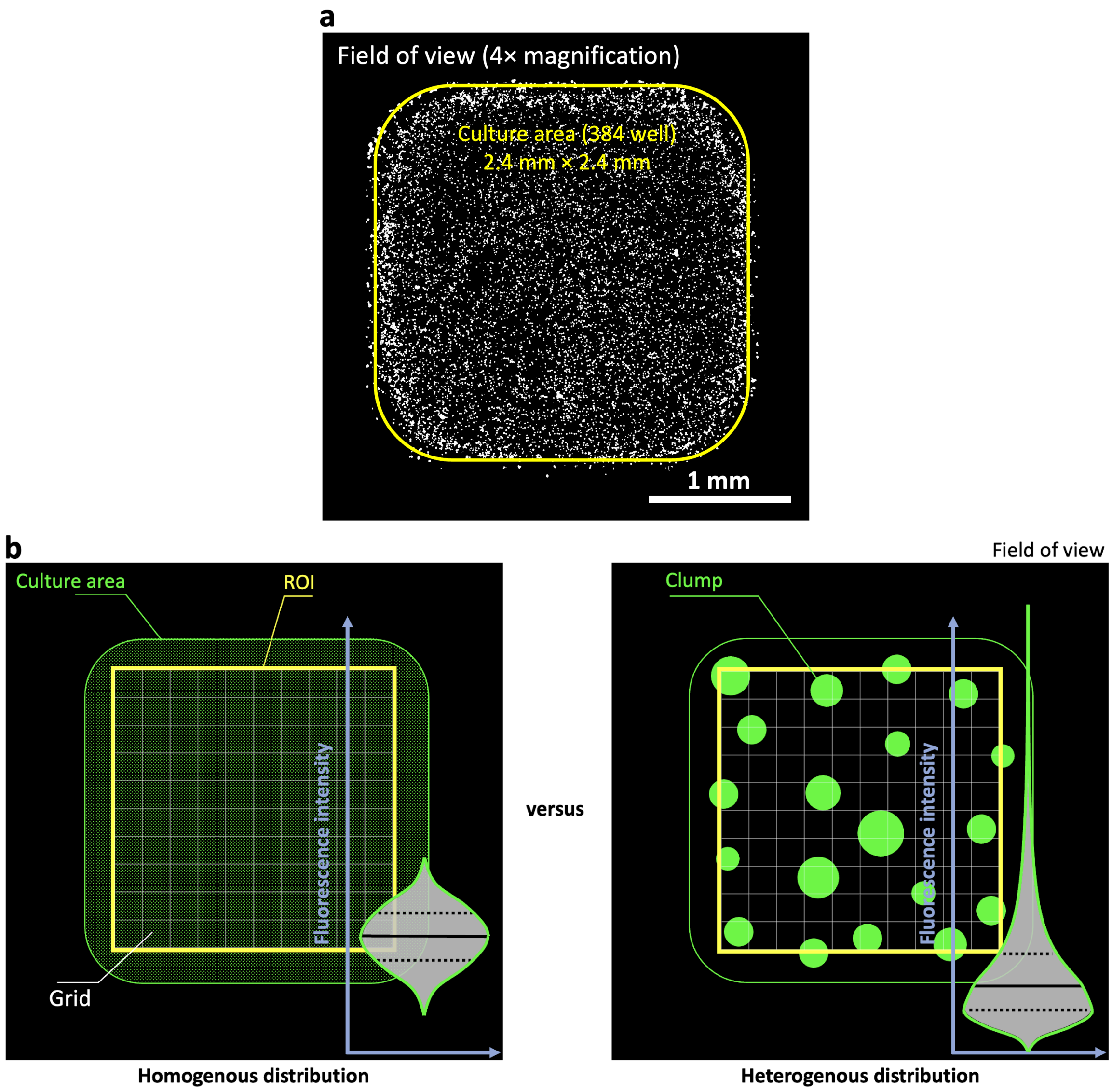
a
Field of view (4× magnification)
Culture area (384 well)
2.4 mm × 2.4 mm
1 mm
b
Field of view
Culture area
ROI
Fluorescence intensity
Grid
Homogenous distribution
versus
Clump
Fluorescence intensity
Heterogenous distribution

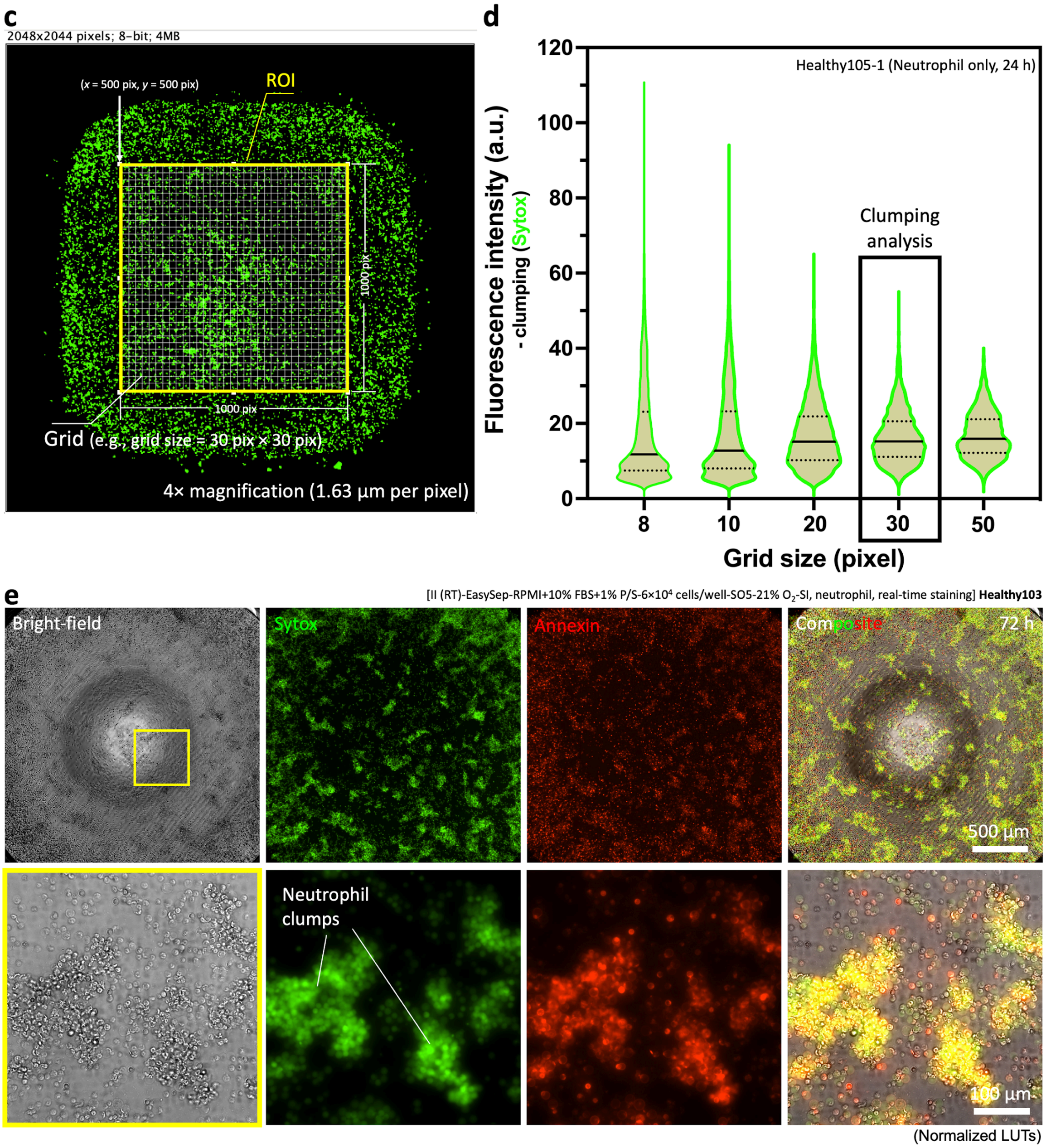


**Supplementary Fig. 8.** Image analysis methods for quantifying neutrophil activation. a) Area fraction analysis. Activation rate (%) = ($S_{\text{particles-Sytox+}}/S_{\text{culture area}}$) × 100% = Area fraction (Image J) × ($S_{\text{field of view}}/S_{\text{culture area}}$) × 100%, where Area fraction (Image J) = $S_{\text{particles-Sytox+}}/S_{\text{field of view}}$, $S$ is for surface area. b) Comparison between homogeneous and heterogeneous (i.e., clumping) particle distribution. The algorithm exploits the fact by dividing a region of interest (ROI) on an image with a grid of squares. The grid size (i.e., the surface area of each square) is defined by the average size of clumps. In homogeneous distribution (i.e., with little clumping), the intensity distribution is visualized as a narrow and symmetrical box-violin band. In comparison, clumping leads to a more polarized and asymmetrical box-violin band. c) A microscopic image in 4× magnification, 2048 × 2044 pixels with activated neutrophils (Sytox+) showing mild clumps. The ROI is 1000 × 1000 pixels. The grid size is 30 × 30 pixels, i.e., Grid30. d) A graph showing evolution of symmetry of the clumping box-violin band as a function of grid size. The larger the grid size, the more homogeneous particle distribution will be. In this work, we used Grid30 to identify the typical clumps formed by activated neutrophils in the standard 2D monoculture. e) Close-up images of neutrophil clumps from experiments.

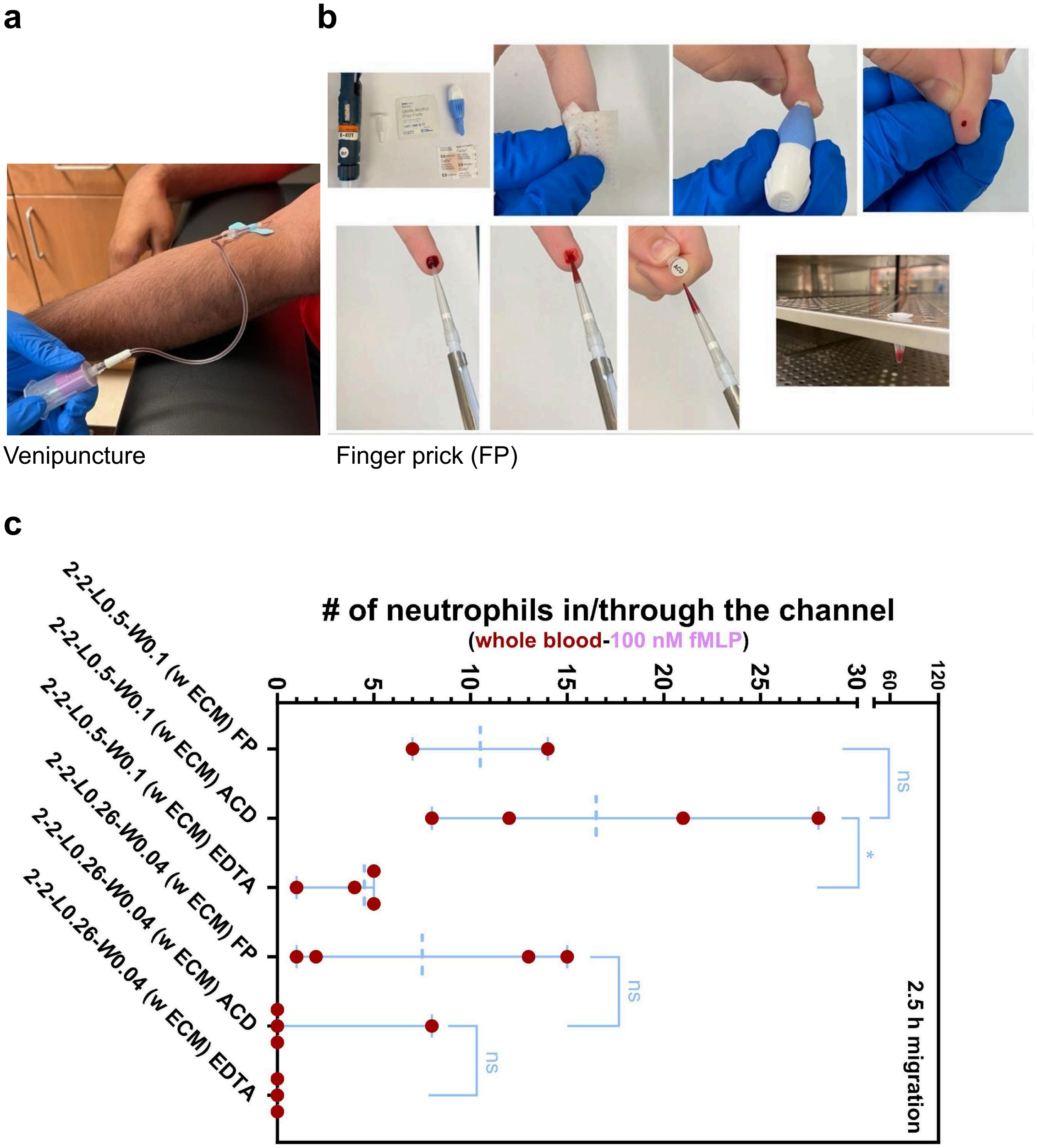


**Supplementary Fig. 9.** Neutrophil recruitment in µ-Blood with whole blood collected from venipuncture and finger prick. a) A camera photo of venipuncture whole blood collection. Different blood tubes (e.g., EDTA - 1.8 mg/mL whole blood, ACD-A - 1:10 dilution with whole blood) were used (Methods). b) Camera photos of finger prick (FP) whole blood collection. The FP blood can be used directly without anticoagulant in 5 min after blood collection (otherwise clotted in the tube) or mixed with 1:10 ACD-A solution and stored in a standard $CO_2$ incubator (37 °C, 18.6% $O_2$, 5% $CO_2$, 95% RH) before use. c, Comparison of FP blood (without anticoagulant), venipuncture blood (with ACD), and venipuncture blood (with EDTA) for neutrophil recruitment against fMLP (100 nM) in μ-Blood. The standard blood collection concentration of EDTA (1.8 mg/mL) strongly inhibits neutrophil migration against 100 nM fMLP (Supplementary Movie 1). Neutrophils respond similarly to fMLP with ACD whole blood and anticoagulant-free FP blood. Error bars are mean ± S.D.. *$P \leq 0.05$, **$P \leq 0.01$, ***$P \leq 0.001$, and ****$P \leq 0.0001$. “ns” represents “not significant”.

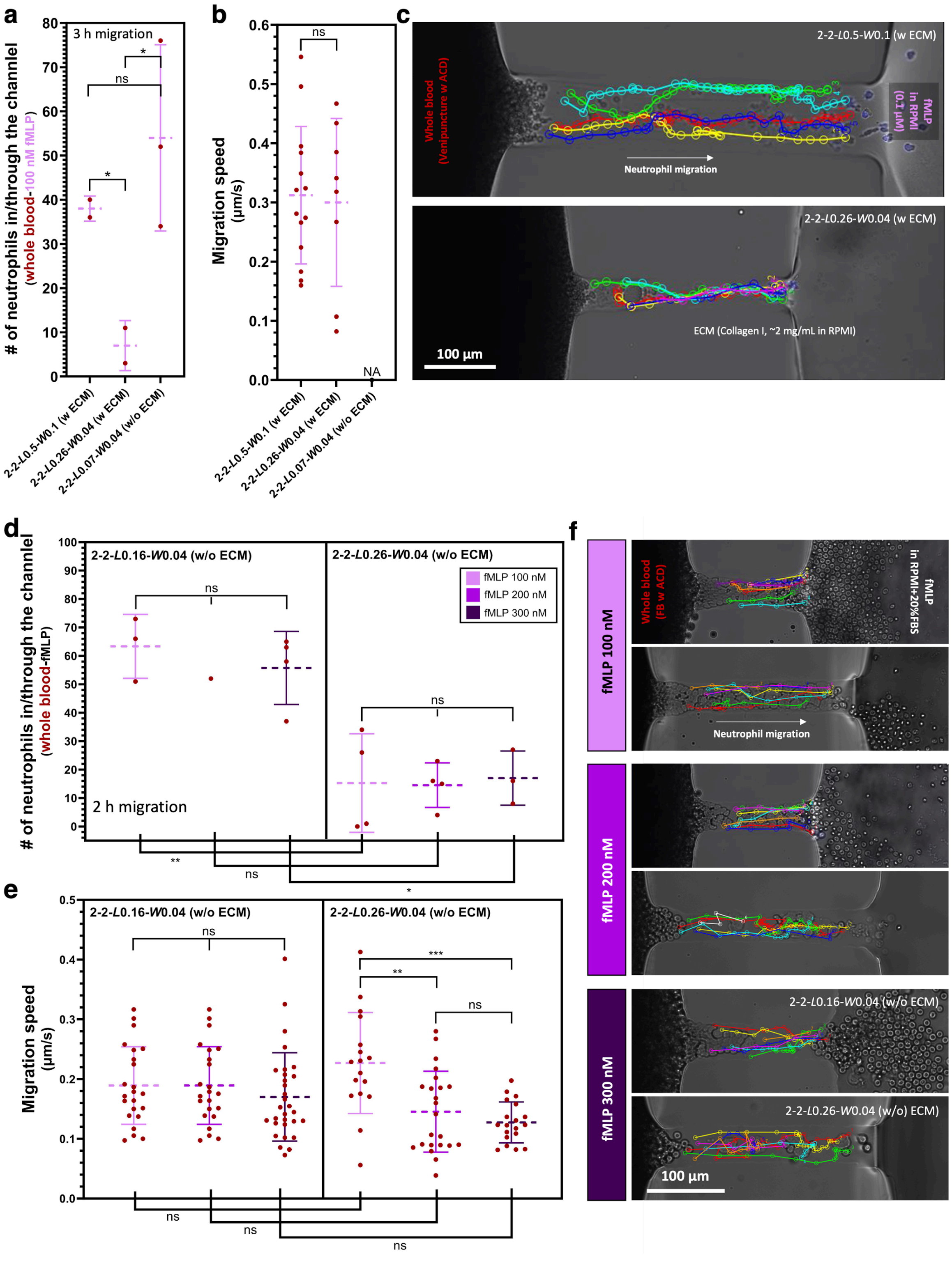
a
3 h migration
### of neutrophils in/through the channel (whole blood-100 nM fMLP)
2-2-L0.5-W0.1 (w ECM)
2-2-L0.26-W0.04 (w ECM)
2-2-L0.07-W0.04 (w/o ECM)
b
Migration speed (μm/s)
NA
c
2-2-L0.5-W0.1 (w ECM)
Whole blood (Venipuncture w ACD)
fMLP in RPMI (0.1 μM)
Neutrophil migration
2-2-L0.26-W0.04 (w ECM)
ECM (Collagen I, ~2 mg/mL in RPMI)
100 μm
d
2-2-L0.16-W0.04 (w/o ECM)
2-2-L0.26-W0.04 (w/o ECM)
fMLP 100 nM
fMLP 200 nM
fMLP 300 nM
2 h migration
### of neutrophils in/through the channel (whole blood-fMLP)
e
Migration speed (μm/s)
f
Whole blood (FB w ACD)
fMLP in RPMI+20%FBS
Neutrophil migration
2-2-L0.16-W0.04 (w/o ECM)
2-2-L0.26-W0.04 (w/o) ECM)
100 μm

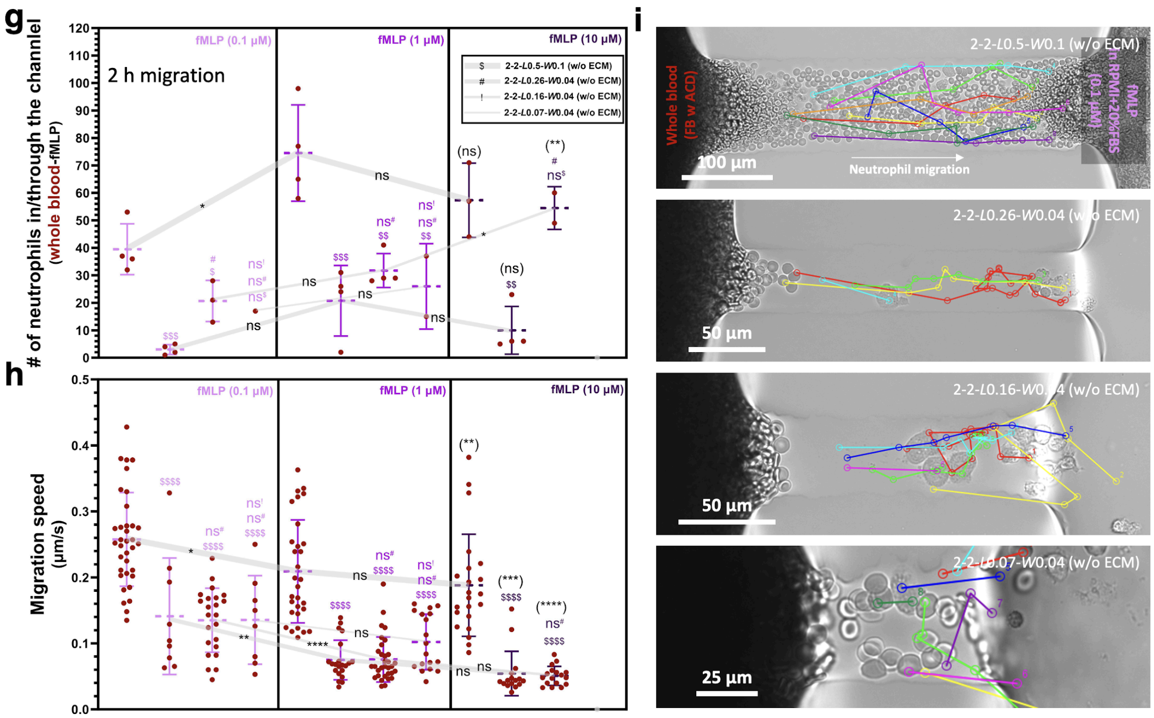


**Supplementary Fig. 10.** Neutrophil recruitment through under-oil microchannels with different dimensions (i.e., channel length and width), with or without ECM coating, and against different fMLP concentrations. a-c) Microchannels w and w/o ECM (100 nM fMLP) (Supplementary Movie 2-1). d-f) Microchannels w/o ECM (100 nM, 200 nM, and 300 nM fMLP) (Supplementary Movie 2-2). g-i) Microchannels w/o ECM (0.1 μM, 1 μM, and 10 μM fMLP) (Supplementary Movie 2-3). Error bars are mean ± S.D.. *$P \le 0.05$, **$P \le 0.01$, ***$P \le 0.001$, and ****$P \le 0.0001$. "ns" represents "not significant".

**Supplementary Table. 1.** Calculation of fluid volume in the microchannels right after sweep.

| I - Right after sweep (Supplementary Fig. 2a) | | | | |
|---|---|---|---|---|
| $W_{channel}$ (μm)* | $H_{channel}$ ($H_{channel}/W_{channel}$ = 1/13) (μm) | $L_{channel}$ (μm)* | Cross-section (μm$^2$) | $V_{channel}$ (pL) |
| 30 | 2.3 | 300 | 46.4 | 13.9 |
| 100 | 7.7 | 500 | 515.2 | 257.6 |
| 200 | 15.4 | 500 | 2061.0 | 1030.5 |
| 300 | 23.1 | 500 | 4637.1 | 2318.6 |
| 500 | 38.5 | 500 | 12881.0 | 6440.5 |

*The channel width and length in this table used for fluid volume calculation are the design values on the photomask of the microchannels. The patterns on the PDMS stamps are 10-20 μm enlarged compared to the design values on the photomask due to the resolution limit of our UV exposure system in photolithography[2], which leads to 10-20 μm increase in channel width and 20-40 μm (doubled due to the channel connecting two spots) decrease in channel length. The channel dimensions in Supplementary Fig. 2c,d) are the measured values on a microscope.

**Supplementary Movie 1.** Influence of anticoagulant (finger prick versus ACD, EDTA) on neutrophil migration in μ-Blood - under-oil microchannels 2-2-*L*0.5-*W*0.1 (w ECM), single-oil (silicone oil 5 cSt) overlay, fMLP in RPMI 100 nM, 2.5 h migration, 900× speed.

**Supplementary Movie 2-1.** Influence of channel dimensions on neutrophil migration in μ-Blood - under-oil microchannels 2-2-*L*0.5-*W*0.1 (w ECM), 2-2-*L*0.26-*W*0.04 (w ECM), and 2-2-*L*0.07-*W*0.04 (w/o ECM), single-oil (silicone oil 5 cSt) overlay, fMLP in RPMI (*L*0.5, *L*0.26) or A-PFP (*L*0.07) 100 nM, 3 h migration, 600× speed.

**Supplementary Movie 2-2.** Influence of channel dimensions and fMLP concentrations on neutrophil migration in μ-Blood - under-oil microchannels 2-2-*L*0.16-*W*0.04 (w/o ECM) and 2-2-*L*0.26-*W*0.04 (w/o ECM), double-oil (silicone oil 20 cSt+Fluorinert FC-40) overlay, fMLP in RPMI + 20% FBS 100 nM, 200 nM and 300 nM, 2 h migration, 600× speed.

**Supplementary Movie 2-3.** Influence of channel dimensions and fMLP concentrations on neutrophil migration in μ-Blood - under-oil microchannels 2-2-*L*0.5-*W*0.1 (w/o ECM), 2-2-*L*0.26-*W*0.04 (w/o ECM), 2-2-*L*0.16-*W*0.04 (w/o ECM), and 2-2-*L*0.07-*W*0.04 (w/o ECM), double-oil (silicone oil 20 cSt+Fluorinert FC-40) overlay, fMLP in RPMI + 20% FBS 0.1 μM, 1 μM, and 10 μM, 2 h migration, 600× speed.

**Supplementary Movie 3.** Healthy122 versus Prostate873 neutrophil migration in μ-Blood (3 h) - under-oil microchannels 2-2-*L*0.26-*W*0.04 (w/o ECM), double-oil (silicone oil 20 cSt+Fluorinert FC-40) overlay, fMLP in A-PFP 100 nM, 1-3 h migration, 360× speed.

**Supplementary Movie 4.** Healthy109 versus Prostate624 neutrophil migration in μ-Blood (1.6 h) - under-oil microchannels 2-2-*L*0.5-*W*0.1 (w/o ECM), double-oil (silicone oil 20 cSt+Fluorinert FC-40) overlay, GFP-*S. aureus* in MHB 5 × $10^8$ CFU/mL, 1.6 h migration, 900× speed.

**Supplementary Movie 5.** Healthy109 versus Prostate624 neutrophil migration in μ-Blood (6 days) - under-oil microchannels 2-2-*L*0.26-*W*0.04 (w/o ECM), double-oil (silicone oil 20 cSt+Fluorinert FC-40) overlay, fMLP in A-PFP 100 nM, 2 h migration, 600× speed.
